# Quantitative LC-MS study of compounds found predictive of COVID-19 severity and outcome

**DOI:** 10.1101/2023.03.17.23287401

**Authors:** Ivayla Roberts, Marina Wright Muelas, Joseph M. Taylor, Andrew S. Davison, Catherine L. Winder, Royston Goodacre, Douglas B. Kell

## Abstract

**INTRODUCTION:** Since the beginning of the SARS-CoV-2 pandemic in December 2019 multiple metabolomics studies have proposed predictive biomarkers of infection severity and outcome. Whilst some trends have emerged, the findings remain intangible and uninformative when it comes to new patients.

**OBJECTIVES:** In this study, we accurately quantitate a subset of compounds in patient serum that were found predictive of severity and outcome.

**METHODS:** A targeted LC-MS method was used in 46 control and 95 acute COVID-19 patient samples to quantitate the selected metabolites. These compounds included tryptophan and its degradation products kynurenine and kynurenic acid (reflective of immune response), butyrylcarnitine and its isomer (reflective of energy metabolism) and finally 3’,4’-didehydro-3’-deoxycytidine, a deoxycytidine analogue, (reflective of host viral defence response). We subsequently examine changes in those markers by disease severity and outcome relative to those of control patients’ levels.

**RESULTS & CONCLUSION:** Finally, we demonstrate the added value of the kynurenic acid / tryptophan ratio for severity and outcome prediction and highlight the viral detection potential of ddhC.

## 1 Introduction

The SARS-CoV-2 virus outbreak which started at end of 2019 in Wuhan, China rapidly transformed into a worldwide pandemic. As of Feb 1^st^ 2023 there have been 753 million confirmed COVID-19 cases with 6.8 million deaths (WHO, 2023b). Although approximately 13 billion vaccines have been administered (WHO, 2023b) the challenge of curbing the pandemic continue due to the international spread and growing list of variants (WHO, 2023a). In response a wave of research was published ranging from understanding the viral origins and molecular composition to societal and economic impact (Else, 2020; Ioannidis et al., 2021).

A number of omics studies investigating disease severity and/or outcome (Costanzo et al., 2022; Mussap and Fanos, 2021) were published that are of specific interest here. When it comes to COVID-19 severity and outcome prediction broad omics data have been shown to improve performance over routinely collected clinical data (López-Hernández et al., 2021). More importantly omics investigations also provided mechanistic insight about the infection. Metabolomics, the subset of omics concerned with small molecules, necessarily amplifies changes in the proteome (Raamsdonk et al., 2001), and has been shown to be a highly sensitive indicator of biochemical differences (Kell and Oliver, 2016) thus is our measurement modality of choice.

Some key findings from published metabolomic investigations of COVID-19 outcome and severity include: the upregulation of the tryptophan degradation pathway (Costanzo et al., 2022; Diray-Arce et al., 2020; Mussap and Fanos, 2021), high levels of some amino acids, and participants in purine & pyrimidine metabolism (Costanzo et al., 2022) including some with specific mention of cytosine. Finally, energy metabolism is also frequently reported (Costanzo et al., 2022) with references to acylcarnitines as in (Dei Cas et al., 2021).

To take these promising findings forward, ultimately to clinical practice, careful validation and quantification studies are necessary. However, to date, very few studies report quantitative data (Karu et al., 2022; López-Hernández et al., 2021; Song et al., 2020; Thomas et al., 2020) and those that did took a broad-spectrum approach to quantification. Furthermore, compound concentrations are difficult to access.

Here we present a quantitative metabolomic study using Liquid Chromatography Mass Spectrometry (LC-MS) on compounds found predictive of disease severity and outcome in our previous untargeted LC-MS discovery study (Roberts et al., 2022). These include tryptophan (TRP), kynurenine (KYN) and kynurenic acid (KYNA) encompassing the TRP degradation pathway. Butyrylcarnitine (C4-carnitine) and its isomeric form (iso-C4-carnitine) were included to represent the acylcarnitine class linked to energy metabolism changes in severe COVID-19 patients. Finally, cytidine and 3’,4’-didehydro-3’-deoxycytidine (ddhC) were included as significant in pyrimidine pathway activation in viral infection.

While our untargeted discovery paper (Roberts et al., 2022) refers to deoxycytidine being measured by its in-source fragmentation to cytosine, further investigation revealed that the fragment was actually breaking from ddhC; an analogue of deoxycytidine produced by the host as a defence mechanism to viral infections (Gizzi et al., 2018). A detailed identification discussion with MS2 fragmentation of ddhC is included in the supplementary information (SI ddhC identification section).

The cohort for this study included 95 COVID-19 patients: a subset of the cohort in the untargeted discovery work (Roberts et al., 2022) constrained by remaining material, but included an additional 46 control patients. The study design aims to investigate accurate metabolite level changes in infection and their added value over routinely collected clinical data. At that end a targeted LC-MS method was developed to allow the accurate and simultaneous measurement of this small number of compounds offering higher sensitivity and accuracy. Our ultimate aim is to support translation of this promising strand of research into clinical practice.

## 2 Results

Overall, analysis of the quantitative measurements of the relevant compounds had good predictive accuracy and the directionality and statistical significance was consistent with our previous discovery study (Roberts et al., 2022). This analysis is expanded upon in the sections below along with additional comparison to the control cohort and selection of clinical data.

The cohort, summarised in Table 1, comprised 141 patients at the Royal Liverpool University Hospital (RLUH) of which 46 were controls and 95 were COVID-19 positive. The 95 SARS-CoV-2 positive patients are a subset the previous discovery and validation cohort (Roberts et al., 2022), where sufficient sample material remained. The COVID-19 patients encompassed 28 mild, 23 intermediate and 44 severe cases with 23 subsequently deceased patients from the severe cases group. Severe cases were defined based on required fraction of inspired oxygen (FIO_2_) > 40% and/or required Continuous Positive Airway Pressure (CPAP) and/or required invasive ventilation and/or did not survive. Intermediate cases required respiratory support, but not to the extent of severe patients and mild cases did not require any respiratory support.

**Table 1.**
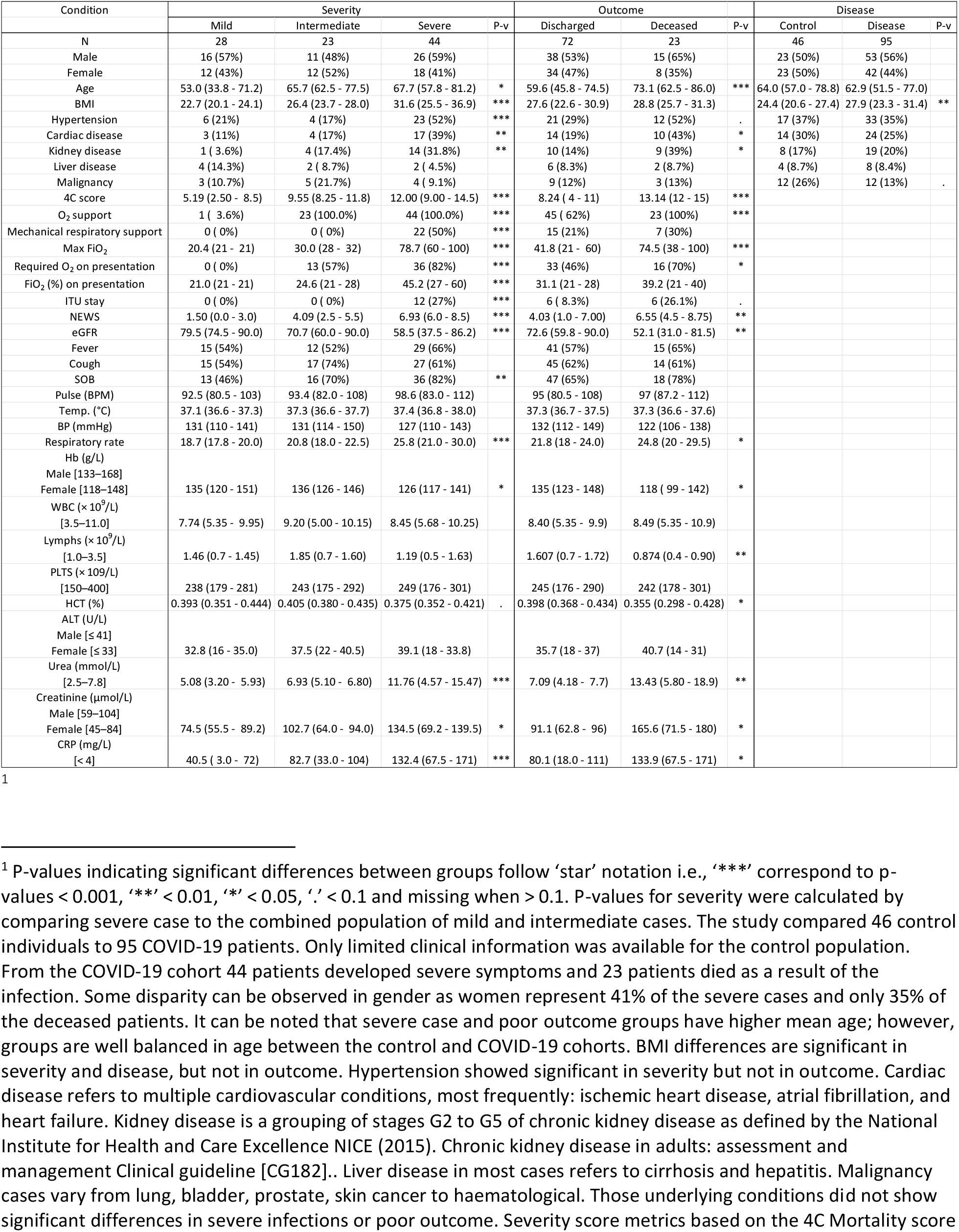
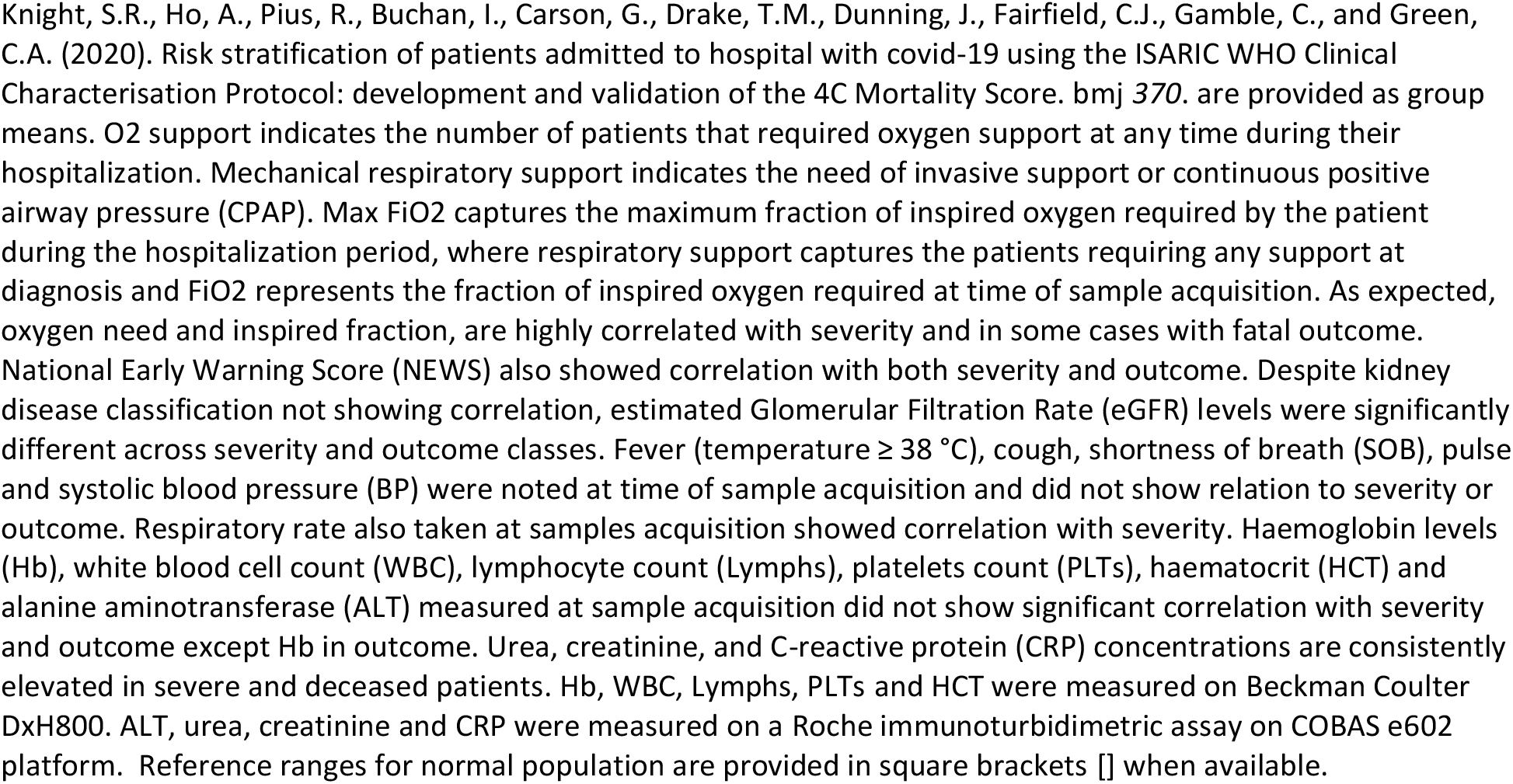
Cohort demographics are presented as counts and percentages (%) for categorical data and means with interquartile range for continuous data.

| Condition | Severity |  |  |  | Outcome |  |  | Disease |  |  |
| --- | --- | --- | --- | --- | --- | --- | --- | --- | --- | --- |
|  | Mild | Intermediate | Severe | P-v | Discharged | Deceased | P-v | Control | Disease | P-v |
| N | 28 | 23 | 44 |  | 72 | 23 |  | 46 | 95 |  |
| Male | 16 (57%) | 11 (48%) | 26 (59%) |  | 38 (53%) | 15 (65%) |  | 23 (50%) | 53 (56%) |  |
| Female | 12 (43%) | 12 (52%) | 18 (41%) |  | 34 (47%) | 8 (35%) |  | 23 (50%) | 42 (44%) |  |
| Age | 53.0 (33.8 - 71.2) | 65.7 (62.5 - 77.5) | 67.7 (57.8 - 81.2) | * | 59.6 (45.8 - 74.5) | 73.1 (62.5 - 86.0) | *** | 64.0 (57.0 - 78.8) | 62.9 (51.5 - 77.0) |  |
| BMI | 22.7 (20.1 - 24.1) | 26.4 (23.7 - 28.0) | 31.6 (25.5 - 36.9) | *** | 27.6 (22.6 - 30.9) | 28.8 (25.7 - 31.3) |  | 24.4 (20.6 - 27.4) | 27.9 (23.3 - 31.4) | ** |
| Hypertension | 6 (21%) | 4 (17%) | 23 (52%) | *** | 21 (29%) | 12 (52%) | . | 17 (37%) | 33 (35%) |  |
| Cardiac disease | 3 (11%) | 4 (17%) | 17 (39%) | ** | 14 (19%) | 10 (43%) | * | 14 (30%) | 24 (25%) |  |
| Kidney disease | 1 (3.6%) | 4 (17.4%) | 14 (31.8%) | ** | 10 (14%) | 9 (39%) | * | 8 (17%) | 19 (20%) |  |
| Liver disease | 4 (14.3%) | 2 (8.7%) | 2 (4.5%) |  | 6 (8.3%) | 2 (8.7%) |  | 4 (8.7%) | 8 (8.4%) |  |
| Malignancy | 3 (10.7%) | 5 (21.7%) | 4 (9.1%) |  | 9 (12%) | 3 (13%) |  | 12 (26%) | 12 (13%) | . |
| 4C score | 5.19 (2.50 - 8.5) | 9.55 (8.25 - 11.8) | 12.00 (9.00 - 14.5) | *** | 8.24 (4 - 11) | 13.14 (12 - 15) | *** |  |  |  |
| O<sub>2</sub> support | 1 (3.6%) | 23 (100.0%) | 44 (100.0%) | *** | 45 (62%) | 23 (100%) | *** |  |  |  |
| Mechanical respiratory support | 0 (0%) | 0 (0%) | 22 (50%) | *** | 15 (21%) | 7 (30%) |  |  |  |  |
| Max FIO<sub>2</sub> | 20.4 (21 - 21) | 30.0 (28 - 32) | 78.7 (60 - 100) | *** | 41.8 (21 - 60) | 74.5 (38 - 100) | *** |  |  |  |
| Required O<sub>2</sub> on presentation | 0 (0%) | 13 (57%) | 36 (82%) | *** | 33 (46%) | 16 (70%) | * |  |  |  |
| FIO<sub>2</sub> (%) on presentation | 21.0 (21 - 21) | 24.6 (21 - 28) | 45.2 (27 - 60) | *** | 31.1 (21 - 28) | 39.2 (21 - 40) |  |  |  |  |
| ITU stay | 0 (0%) | 0 (0%) | 12 (27%) | *** | 6 (8.3%) | 6 (26.1%) | . |  |  |  |
| NEWS | 1.50 (0.0 - 3.0) | 4.09 (2.5 - 5.5) | 6.93 (6.0 - 8.5) | *** | 4.03 (1.0 - 7.00) | 6.55 (4.5 - 8.75) | ** |  |  |  |
| eGFR | 79.5 (74.5 - 90.0) | 70.7 (60.0 - 90.0) | 58.5 (37.5 - 86.2) | *** | 72.6 (59.8 - 90.0) | 52.1 (31.0 - 81.5) | ** |  |  |  |
| Fever | 15 (54%) | 12 (52%) | 29 (66%) |  | 41 (57%) | 15 (65%) |  |  |  |  |
| Cough | 15 (54%) | 17 (74%) | 27 (61%) |  | 45 (62%) | 14 (61%) |  |  |  |  |
| SOB | 13 (46%) | 16 (70%) | 36 (82%) | ** | 47 (65%) | 18 (78%) |  |  |  |  |
| Pulse (BPM) | 92.5 (80.5 - 103) | 93.4 (82.0 - 108) | 98.6 (83.0 - 112) |  | 95 (80.5 - 108) | 97 (87.2 - 112) |  |  |  |  |
| Temp. (°C) | 37.1 (36.6 - 37.3) | 37.3 (36.6 - 37.7) | 37.4 (36.8 - 38.0) |  | 37.3 (36.7 - 37.5) | 37.3 (36.6 - 37.6) |  |  |  |  |
| BP (mmHg) | 131 (110 - 141) | 131 (114 - 150) | 127 (110 - 143) |  | 132 (112 - 149) | 122 (106 - 138) |  |  |  |  |
| Respiratory rate | 18.7 (17.8 - 20.0) | 20.8 (18.0 - 22.5) | 25.8 (21.0 - 30.0) | *** | 21.8 (18 - 24.0) | 24.8 (20 - 29.5) | * |  |  |  |
| Hb (g/L) |  |  |  |  |  |  |  |  |  |  |
| Male [133–168] |  |  |  |  |  |  |  |  |  |  |
| Female [118–148] | 135 (120 - 151) | 136 (126 - 146) | 126 (117 - 141) | * | 135 (123 - 148) | 118 (99 - 142) | * |  |  |  |
| WBC (× 10<sup>9</sup>/L) |  |  |  |  |  |  |  |  |  |  |
| [3.5–11.0] | 7.74 (5.35 - 9.95) | 9.20 (5.00 - 10.15) | 8.45 (5.68 - 10.25) |  | 8.40 (5.35 - 9.9) | 8.49 (5.35 - 10.9) |  |  |  |  |
| Lymphs (× 10<sup>9</sup>/L) |  |  |  |  |  |  |  |  |  |  |
| [1.0–3.5] | 1.46 (0.7 - 1.45) | 1.85 (0.7 - 1.60) | 1.19 (0.5 - 1.63) |  | 1.607 (0.7 - 1.72) | 0.874 (0.4 - 0.90) | ** |  |  |  |
| PLTS (× 10<sup>9</sup>/L) |  |  |  |  |  |  |  |  |  |  |
| [150–400] | 238 (179 - 281) | 243 (175 - 292) | 249 (176 - 301) |  | 245 (176 - 290) | 242 (178 - 301) |  |  |  |  |
| HCT (%) | 0.393 (0.351 - 0.444) | 0.405 (0.380 - 0.435) | 0.375 (0.352 - 0.421) | . | 0.398 (0.368 - 0.434) | 0.355 (0.298 - 0.428) | * |  |  |  |
| ALT (U/L) |  |  |  |  |  |  |  |  |  |  |
| Male [≤ 41] |  |  |  |  |  |  |  |  |  |  |
| Female [≤ 33] | 32.8 (16 - 35.0) | 37.5 (22 - 40.5) | 39.1 (18 - 33.8) |  | 35.7 (18 - 37) | 40.7 (14 - 31) |  |  |  |  |
| Urea (mmol/L) |  |  |  |  |  |  |  |  |  |  |
| [2.5–7.8] | 5.08 (3.20 - 5.93) | 6.93 (5.10 - 6.80) | 11.76 (4.57 - 15.47) | *** | 7.09 (4.18 - 7.7) | 13.43 (5.80 - 18.9) | ** |  |  |  |
| Creatinine (μmol/L) |  |  |  |  |  |  |  |  |  |  |
| Male [59–104] |  |  |  |  |  |  |  |  |  |  |
| Female [45–84] | 74.5 (55.5 - 89.2) | 102.7 (64.0 - 94.0) | 134.5 (69.2 - 139.5) | * | 91.1 (62.8 - 96) | 165.6 (71.5 - 180) | * |  |  |  |
| CRP (mg/L) |  |  |  |  |  |  |  |  |  |  |
| [< 4] | 40.5 (3.0 - 72) | 82.7 (33.0 - 104) | 132.4 (67.5 - 171) | *** | 80.1 (18.0 - 111) | 133.9 (67.5 - 171) | * |  |  |  |

COVID-19 patient serum was acquired at hospital admission in 2 batches: the first a batch of 31 patients early in the pandemic (April – June 2020) and the second a batch of 64 patients in early 2021. Control samples were drawn in (July – October 2020).

The quality of the quantitative results was validated based on calibration curve linearity, accuracy, precision, any matrix effects, and reproducibility following FDA guidelines described in (FDA, 2018). Detailed results on data quality assessment are presented in the supplementary information section 3.1.

Quantitated serum concentration ranges are presented in Table 2. Compound concentrations per group did not follow a normal distribution; thus, the quantile ranges are described in the format of median (Q2 – Q3), where median represents 50^th^ percentile, Q2 the 25^th^ percentile and Q3 the 75^th^ percentile. Compound trends and significance in different groups based on statistical analysis is reviewed in detail in the following section.

**Table 2.**
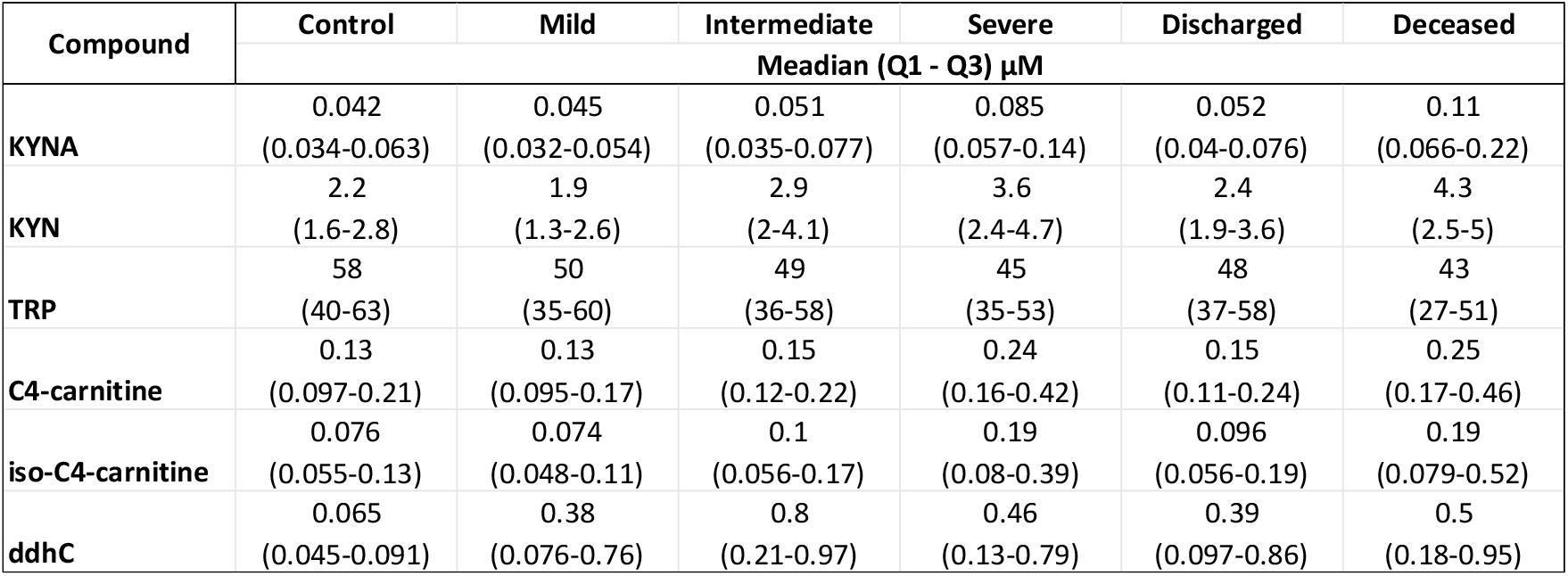
Compound concentrations per group are in μM following the format of Q2 (Q1-Q3). Where Q2 represents the median value of the distribution and Q1 & Q3 are the 25^th &^ 75^th^ percentile respectivly. This format was selected as the measurements per group were not normally distributed.

| Compound | Control | Mild | Intermediate | Severe | Discharged | Deceased |
| --- | --- | --- | --- | --- | --- | --- |
| | Median (Q1 - Q3) $\mu\text{M}$ | | | | | |
| **KYNA** | 0.042<br>(0.034-0.063) | 0.045<br>(0.032-0.054) | 0.051<br>(0.035-0.077) | 0.085<br>(0.057-0.14) | 0.052<br>(0.04-0.076) | 0.11<br>(0.066-0.22) |
| **KYN** | 2.2<br>(1.6-2.8) | 1.9<br>(1.3-2.6) | 2.9<br>(2-4.1) | 3.6<br>(2.4-4.7) | 2.4<br>(1.9-3.6) | 4.3<br>(2.5-5) |
| **TRP** | 58<br>(40-63) | 50<br>(35-60) | 49<br>(36-58) | 45<br>(35-53) | 48<br>(37-58) | 43<br>(27-51) |
| **C4-carnitine** | 0.13<br>(0.097-0.21) | 0.13<br>(0.095-0.17) | 0.15<br>(0.12-0.22) | 0.24<br>(0.16-0.42) | 0.15<br>(0.11-0.24) | 0.25<br>(0.17-0.46) |
| **iso-C4-carnitine** | 0.076<br>(0.055-0.13) | 0.074<br>(0.048-0.11) | 0.1<br>(0.056-0.17) | 0.19<br>(0.08-0.39) | 0.096<br>(0.056-0.19) | 0.19<br>(0.079-0.52) |
| **ddhC** | 0.065<br>(0.045-0.091) | 0.38<br>(0.076-0.76) | 0.8<br>(0.21-0.97) | 0.46<br>(0.13-0.79) | 0.39<br>(0.097-0.86) | 0.5<br>(0.18-0.95) |

### 2.1 Statistical analysis results

Figure 1 and 2 show box and whisker plots for compounds and key ratios by (a) severity and (b) outcome groups. One can see an increase in TRP degradation products and C4-carnitines in severity and outcome. KYNA particularly showed a strong discrimination potential between deceased and discharged patients with KYN and KYNA increasing progressively with severity. On the other hand, TRP decreased with severity. C4-carnitine showed a significant increase in severe cases as opposed to mild and intermediate (Figure 1). Finally, 3’,4’-didehydro-3’-deoxycytidine (ddhC) appeared to be extremely good at discriminating COVID-19 cases *vs*. control; however, its correlation to severity and outcome was not as clear.

**Figure 1.**
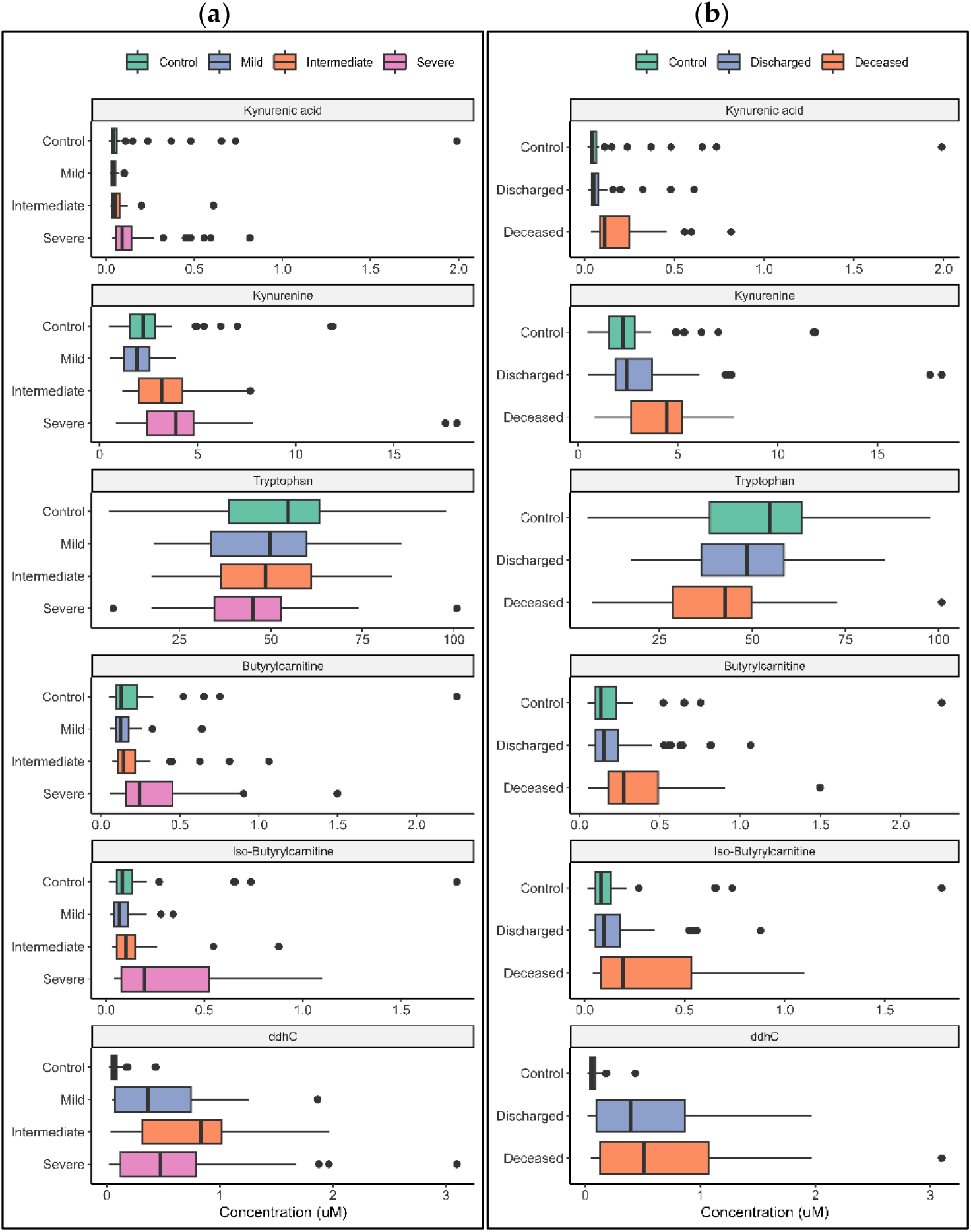
Compound concentrations in COVID-19 infection (a) severity and (b) outcome compared to control patients. Boxes represent the quartiles Q1 to Q3 with Q2 (i.e., median) line in the middle. The ‘whiskers’ depict the upper and lower limit i.e., Q1 ± (Q3-Q1). Outliers are represented in black circles.

**Figure 2.**
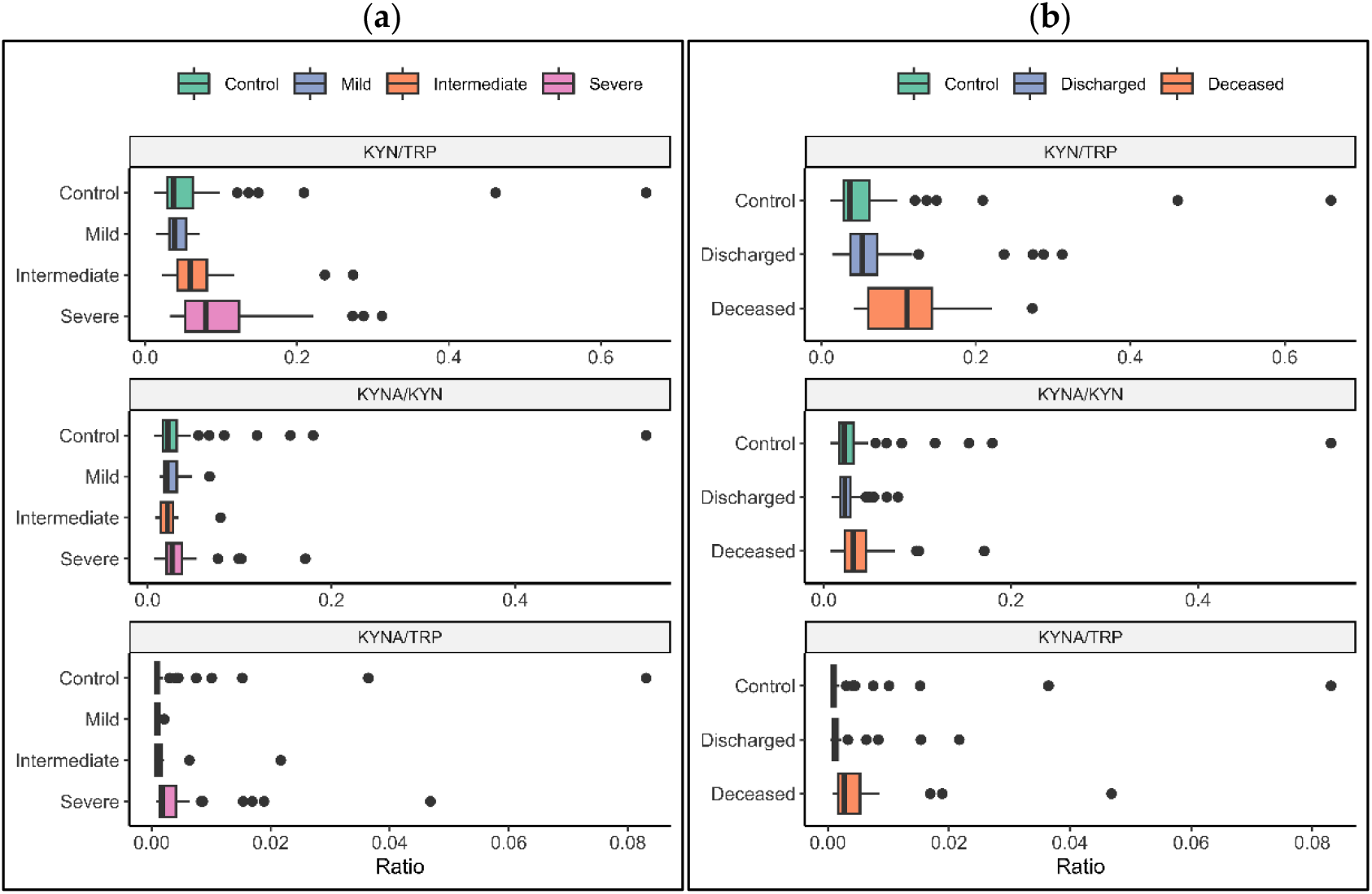
Compound ratios in COVID-19 infection by (a) severity and (b) outcome including control population. Boxes represent the quartiles Q1 to Q3 with Q2 (i.e., median) line in the middle. The ‘whiskers’ depict the upper and lower limit i.e., Q1 ± (Q3-Q1). Outliers are represented in black circles.

In the following sections we discuss in detail the results of logistic regression of individual compounds to SARS-CoV-2 infection, severity, and outcome, a summary of which is shown in Table 3. A more detailed version for the control *vs*. COVID-19 population, mild and discharged groups, can be found in SI Table 7 and for COVID-19 severity and outcome including BMI correction in SI Table 8. Furthermore, we investigated the smaller sub-group of severe COVID-19 cases (*n*=44) and target compound significance in cases of poor outcome (*n*=23) compared to discharged patients. In addition, the commonly used ratios of KYN/TRP, KYNA/TRP and KYNA/KYN were included.

**Table 3.**
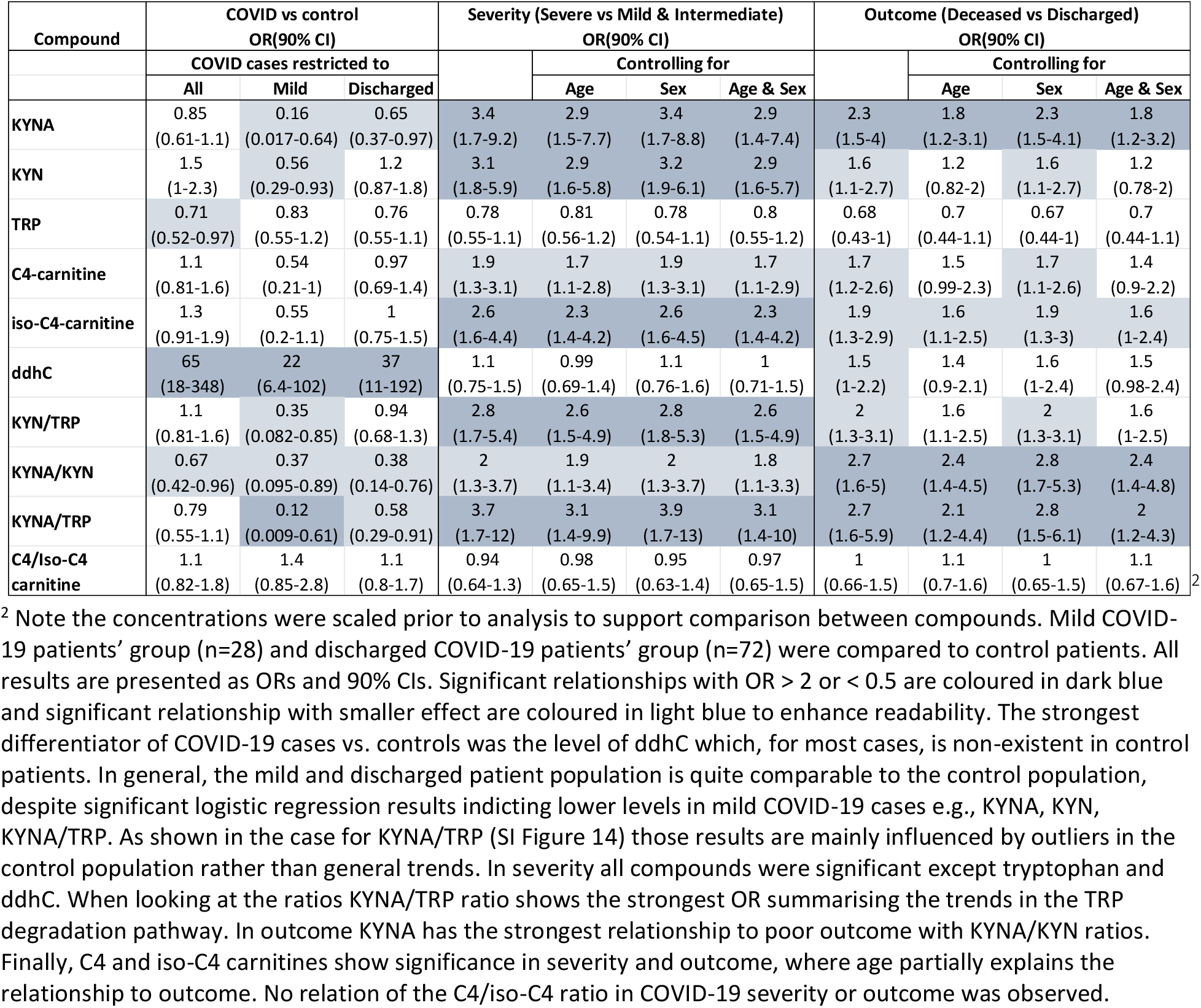
Results from logistic regression comparing COVID-19 to control, severe cases to non-severe and deceased to discharged patients.

### 2.1.1 KYNA, KYN, TRP

KYNA serum concentrations showed the strongest discrimination for both outcome and severity and this was consistent after adjusting for age and sex. KYNA levels appeared significantly lower in mild and discharged patients compared to controls; however, those results are mostly due to the outliers in the control population as demonstrated for the KYNA/TRP ratio in SI figure 14. On the other hand, in severe cases and those with poor outcome KYNA showed one of the strongest OR (Table 3). When corrected for demographic factors KYNA levels appeared to be partially explained by age in both severity and outcome and by BMI in severity (SI Table 8); however, sex did not affect the OR nor the CI. KYN, similarly to KYNA for severity, had a negative relationship between mild COVID-19 cases compared and controls with an OR in the range of 2.8 to 3.2. However, in outcome, KYN changes were not as significant. For TRP, logistic regression indicated tendency for decreasing with severity and outcome but was not significant in any group.

Furthermore, we performed the analysis using common ratios of KYN/TRP, KYNA/TRP and KYNA/KYN. KYN/TRP ratio consistently increased with severity and outcome. Interestingly, in outcome, the significance of KYN/TRP is partially explained by age and BMI as indicated by the drop of OR and CI when corrected. The KYNA/KYN ratio shows higher values in control patients compared to the COVID-19 cohort i.e., OR lower than 1 with CI lower than 1. This is one more time most likely due to the large variance in control patients. However, the KYNA/KYN ratio is strongly linked to outcome and more moderately to severity of COVID-19. This indicates that the increase in KYNA in poor outcome cases is stronger than the increase in KYN compared to severe cases where the increase in KYN reflects more closely the increase in KYNA. Finally, the KYNA/TRP ratio which covers multiple steps of the TRP degradation pathway shows significant increase in both COVID-19 severity and outcome making this ratio the most informative. The difference in COVID-19 vs control population appears to be driven once again by the large variance in the control cohort as shown in the SI Figure 14.

#### 2.1.2 C4-carnitines

No significance was observed in C4 and iso-C4-carnitine when comparing control patients to COVID-19 patients. Negative ORs were found in the case of mild COVID-19 patients, indicating lower level compared to controls, however, our analysis shows this to not be significant.

On the other hand, C4-carnitine showed a significant connection to COVID-19 severity, and this was further amplified in its isomer, iso-C4. The correlation appears to be reduced when correcting for patient age and BMI. Contrary to C4, iso-C4-carnitine relationship to severity appears stronger when corrected for BMI. In outcome, C4-carnitines showed increase with poor outcome, however, the relationship was not as strong as in severity and once again appeared to be explained by age and BMI (SI Table 7 & SI Table 8). Finally, the C4 to iso-C4-carnitine ratio was not found significant in any of the investigated conditions i.e., disease, COVID-19 severity, or outcome.

#### 2.1.3 3’,4’-didehydro-3’-deoxycytidine

The final compound to be investigated, ddhC, showed an excellent ability to discriminate diseased from control patients. ddhC concentrations in control cases tended to be non-existent (< 0.05 µM), close to LLoQ level for this compound (SI Table 4). Despite its strong link to disease when it comes to COVID-19 severity and outcome, ddhC was not as informative (Table 3). In COVID-19 outcome, the tendency was for higher levels in poor outcome however the ORs were highly uncertain and partially explained by age.

**Table 4.**
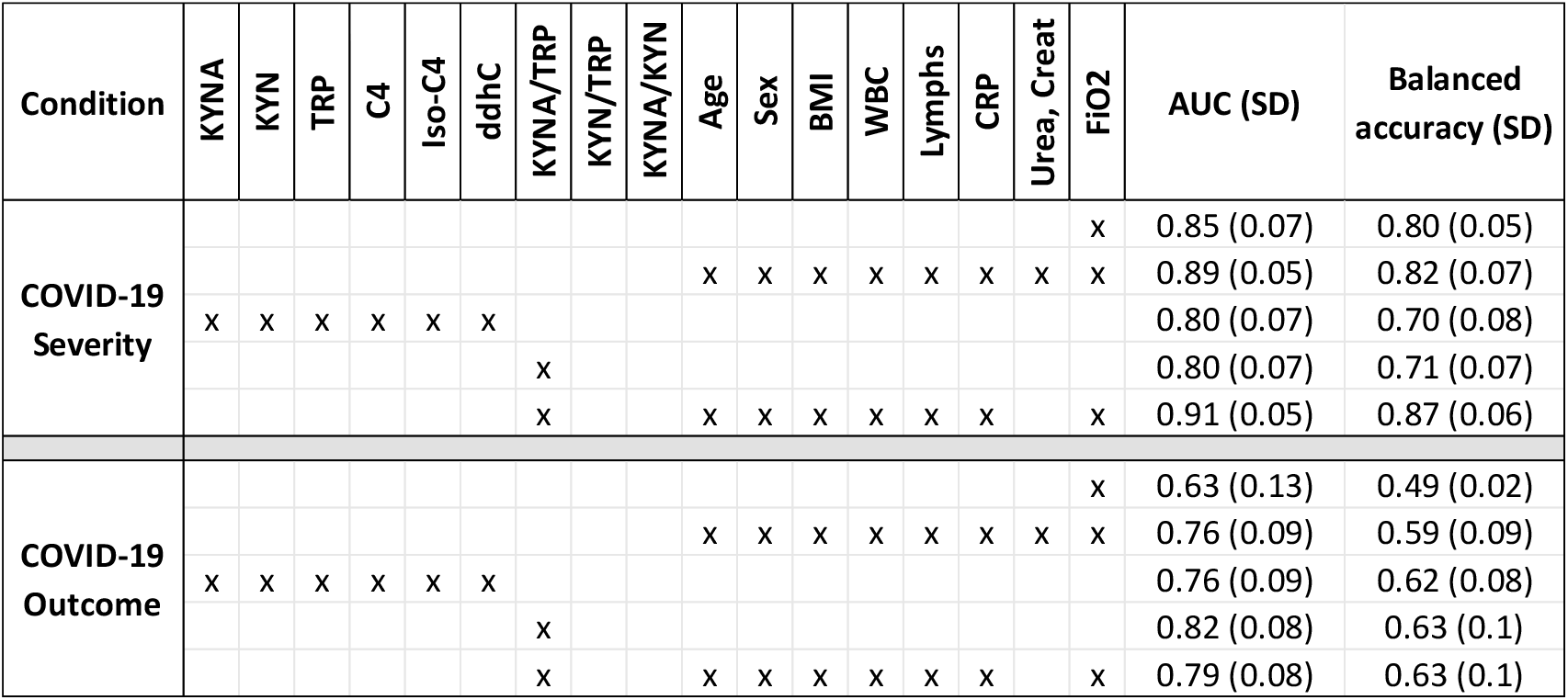
Results from multi compound logistic regression presented in terms of mean area under the curve (AUC) and its standard deviation (SD) as calculated by Monte Carlo cross validation. In addition, mean balanced accuracy is presented to aid interpretation (with threshold of 50%). A model based on existing clinical data predicts severity very well mainly because FIO_2_ is key to determining severity. The same model in COVID-19 outcome provides a reasonable performance. In both cases the addition of KYNA/TRP ratio improves the model and in COVID-19 outcome KYNA/TRP offers more predictive power than clinical data.

#### 2.1.4 Outcome in severe cases

Despite the relatively small sample size of severe COVID-19 cases (*n*=44), we investigated the significance of the targeted compounds in poor outcome (n = 23) compared to discharged severe COVID-19 patients (n = 21). The large CI in the results reflect the small sample size however, a few significant trends emerged as shown in SI Figure 15 & 16 and SI Table 9. Most noticeably, KYNA and KYNA/KYN ratio were significantly elevated in poor COVID-19 outcome. The OR in both cases also fell when corrected for age, indicating some correlation, but remained significant. Moreover, ddhC tended to be higher in poor COVID-19 outcome cases and its correlation to outcome appears to be stronger when corrected for sex, indicating possible gender differences in viral response. Once again, KYNA/TRP ratio showed strong significance and potential to discriminate poor outcome likelihood.

#### 2.1.5 Joint modelling of compounds and clinical measurements

In the final analysis we performed logistic regression on the joint set of compounds both with and without clinical data with the goal of determining overall discriminative power. The clinical data included physiological factors (Age, Sex and BMI) and frequently measured clinical tests (WBC, Lymphs, Urea, Creatinine, CRP and FiO_2_). The results are shown in Table 4.

COVID-19 severity in this cohort was mostly defined by the required oxygen support levels, therefore unsurprisingly FiO_2_ is extremely predictive in severity on its own with an AUC 0.85 (SD 0.07) as shown in Table 4. However, when it comes to COVID-19 outcome, FiO_2_ was not informative. A model including the demographics and clinical data has a very strong predictive power in COVID-19 severity AUC 0.89 (SD 0.5), but also offers information in outcome with 0.76 (SD 0.9).

Interestingly, the KYNA/TRP ratio is as predictive as all 6 metabolites for severity and more predictive in outcome. This shows, with the amount of data in this study, and the correlations between compounds, that this simple ratio is highly informative. Moreover, KYNA/TRP performed better than the clinical and demographics data in outcome.

Building on this finding, we explored the added predictive value of KYNA/TRP to models with clinical and demographic data. Creatinine and urea were removed from this model as they appear strongly correlated to KYNA (see SI Figure 17) but had inferior predictive performance. The addition of KYNA/TRP provided marginal improvements to severity prediction (AUC from 0.89 to 0.91) but for outcome prediction, KYNA/TRP ratio alone remained the best indicator with AUC of 0.82 (SD 0.08).

## 3 Discussion

In this study we accurately measured the concentrations of selected metabolites in control and COVID-19 patient serum samples by LC-MS. The compounds (C4 and iso-C4 carnitine, TRP, KYN, KYNA and ddhC) were selected to reflect the main pathways found predictive of COVID-19 severity and outcome in our previous untargeted LC-MS metabolomics study (Roberts et al., 2022). Upregulation and downregulation trends in our findings are consistent with published studies (Costanzo et al., 2022; Mussap and Fanos, 2021); TRP levels tends to decrease while KYN and KYNA levels increase with COVID-19 severity and poor outcome. We measured KYN increase from 2.2 µM in controls and 1.9 µM in mild infections to 3.6 µM in severe cases, a nearly 2-fold change, and 4.3 µM in poor outcome. Likewise, KYNA concentrations nearly doubled in severe cases (from 0.045 to 0.085 µM) with even stronger increase in deceased patients at 0.11 µM. Equally, we measured C4-carnitine levels increase to nearly 2-fold in severe COVID-19 cases.

### 3.1 Quantitative result differences across studies

There were some differences in the concentration of compounds quantified across different studies. In general, group concentrations of TRP, KYN and C4-carnitine measured by Lopez *et al*. (López-Hernández et al., 2021) are in agreement with our results. However, average concentrations of TRP in Thomas *et al*. (Thomas et al., 2020) appear higher, while Karu *et al*. (Karu et al., 2022) reports lower than ours average TRP levels. KYN levels in the latter two publications (Karu et al., 2022; Thomas et al., 2020) were generally higher than our findings, and KYNA was surprisingly higher in Thomas *et al*. (Thomas et al., 2020). To our knowledge, no studies have reported ddhC concentrations in COVID-19 patients.

These differences could easily be attributed the combination of cohort differences such as demographics and geographical origin but also to quantitative methods differences. Studies discussed previously performed quantitation for numerous metabolites at the same time. In this approach, data accuracy could be impacted by overlapping retention times and lack of dedicated internal standards. Moreover, it is hard to determine optimal sample dilution levels, LC-MS method, and source conditions for hundreds of compounds simultaneously. While developing our method, we found no common sample dilution level that allowed for effective quantification of cytidine (average serum concentration ∼0.2 µM (Wishart et al., 2007)) and tryptophan (average serum concentration 50-80 µM (Wishart et al., 2007)). When TRP was within its linear range, cytidine levels dropped under the LLoQ and when cytidine was within its linear range, TRP levels raised above the ULoQ. Finally, in our method validation we found that KYN suffered from very high (nearly 3-fold) matrix effect between calibration curve levels in replacement matrix (or water) and serum levels as detailed in the SI section 3.1. If not corrected for matrix factor, concentrations will be greatly overestimated, i.e., nearly 3-fold higher levels could be calculated. Those observations demonstrate that more narrowly targeted accurate quantitative measurements are required if we are to enable large, distributed studies and support clinical use of metabolomics findings. It is also possible, that difference in sample storage time, as discussed further in the limitations section, could have resulted in compound degradation and therefore impact quantitative results.

### 3.2 Quantitated compounds biological role

In the following sections we examine each group of compounds in detail. We attempt to put the main observations from this study into the context of known biological processes and propose theories on the compounds’ involvement in COVID-19 severity and outcome. However, it is important to note that no causality can be deduced from this study design therefore all mechanistic hypotheses are speculative at this stage.

#### 3.2.1 KYNA, KYN, TRP

The TRP degradation pathway to KYN and KYNA (Figure 3) is frequently reported as being upregulated in COVID-19 patient plasma, serum (Costanzo et al., 2022; Mussap and Fanos, 2021) and also urine (Dewulf et al., 2022). TRP decreases are often observed but not always significant, perhaps due to naturally high variance across the population. A key motivation for measuring TRP here is to adopt a representation based on KYN/TRP and KYNA/TRP ratios that describe enzymatic activity in the degradation pathway.

**Figure 3.**
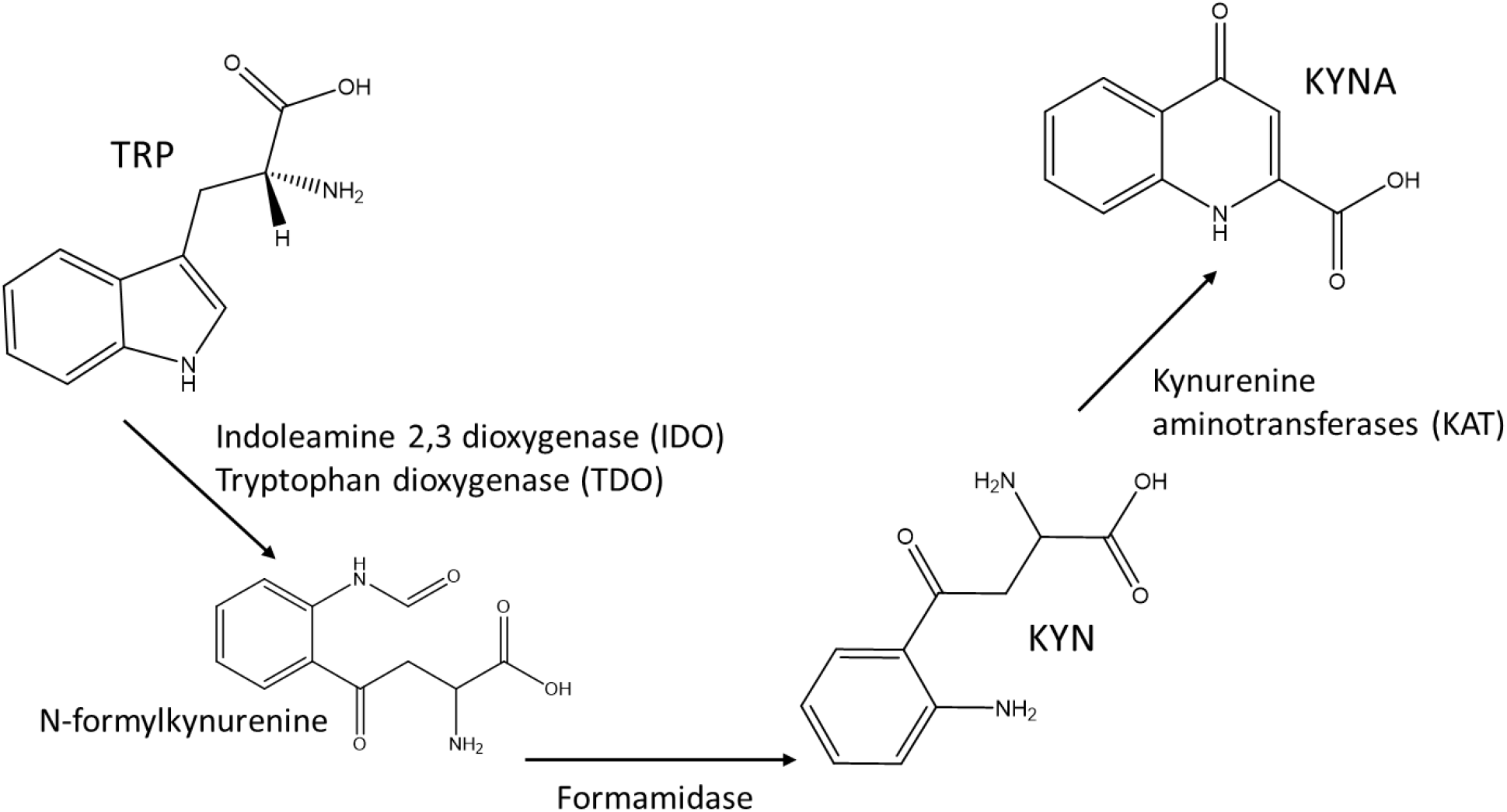
TRP degradation pathway, with associated enzymes.

In our study cohort, KYN/TRP and KYNA/TRP ratios showed the strongest significance in COVID-19 severity (Table 3). For outcome on the other hand, KYNA/KYN ratios provided the best discrimination, showing that in poor outcome the increase in KYNA is stronger than the increase in KYN (Table 3 and SI Table 8). This was further supported by the observation that KYNA was one of the few significant compounds in deceased severe patients compared to discharged severe patients (SI Table 9). Furthermore, when exploring the multi compound predictive models (Table 4), KYNA/TRP ratio improved predictive performance of clinical data in severity models and, in isolation, showed the best performance in outcome.

KYN, and more specifically, KYNA have been most studied for their link to neurodegenerative diseases. While it is generally believed that low levels of KYNA are neuroprotective, higher levels appear to contribute to the symptoms of schizophrenic patients (Kozak et al., 2014; Tanaka et al., 2020). Some attention has been drawn also to the potential role of KYN and KYNA high levels in SARS-CoV-2 infection in neurological symptoms of COVID-19 patients (Collier et al., 2021). However, contrary to its upstream metabolites TRP and KYN, KYNA cannot cross the blood brain barrier. It is therefore more interesting to examine the origin and effects of high KYNA blood levels, and the link to inflammation.

Proinflammatory cytokines upregulate the expression of indoleamine 2,3-dioxygenase (IDO) which subsequently induces an increase in TRP catabolism in the KYN pathway. The resulting higher levels of KYN and its metabolites then have suppressing effects on T-cell proliferation whilst supporting the production of regulatory T-cells (Savitz, 2020). More specifically, KYNA has been found to act as an antioxidant as well as an immunosuppressant (Lugo-Huitrón et al., 2011; Wirthgen et al., 2018). As such, KYNA plays a significant role in protecting from tissues damage in the case of an overactive immune response. In short, while cytokines upregulate the KYN pathway, KYN and KYNA serve as a negative feedback loop creating a more immunotolerant environment.

It has long been known that TRP degradation to KYN is upregulated in HIV, Hepatitis C and herpes simplex, flavivirus (Dengue) viral infections (Collier et al., 2021; Diray-Arce et al., 2020), but also in bacterial infections such as *Mycobacterium tuberculosis* (tuberculosis) (Diray-Arce et al., 2020). Whilst an increase of KYN metabolites is associated with host response to infection, the immunosuppressant effect of KYNA have been suspected to allow immune evasion in tuberculosis (Diray-Arce et al., 2020) and tumour survival in cancer (Walczak et al., 2020). All previously mentioned infections where high ratios of KYN and/or KYNA to TRP have been detected, the occurrence of those hight levels were associated with severe prognoses.

Our observations of an increased TRP/KYN, TRP/KYNA ratio in COVID-19 severity and increase of KYNA/KYN ratio in COVID-19 patients with poor outcome appears to be in line with the hypothesis of immune response suppression by TRP degradation products. Moreover, this hypothesis is supported by the lower lymphocyte count levels in patients with poor outcome as shown in Table 1, however no correlation between KYNA levels and lymphocyte counts was observed in our data, i.e., at hospital admission time as shown in SI Figure 18.

#### 3.2.2 C4-carnitines

C4-carnitine was selected for this quantitative study to represent the acylcarnitines that we found elevated in patients with severe COVID-19 and poor outcome in our discovery study (Roberts et al., 2022). LC separation allowed us to distinguish between C4 and its isomer therefore we quantitated both separately. However, despite varying proportions between individuals, those variations did not corelate with COVID-19 infection or disease severity / outcome as shown by the C4-carnitine/iso-C4 carnitine ratio in Table 3 and SI Table 7, 8 & 9.

While most of the published studies report acylcarnitine levels as increased in COVID-19 patients in relation to severity (Barberis et al., 2020; Castañé et al., 2022; López-Hernández et al., 2021), one study reported a decrease in SARS-CoV-2 infected individuals (Thomas et al., 2020). Possibly this could be compared to the reduction we observe in mild COVID-19 patients compared to controls. However, the C4-carnitine decrease in mild COVID-19 patients failed to prove significant in our data (see SI Table 7). On the other hand, increased of C4 and iso-C4-carnitine with severity were strongly marked in our cohort. Interestingly this increase appeared to be partially explained by age and BMI in C4-carnitine, but not by BMI in its isomeric form as shown in SI Table 8.

In poor outcome COVID-19 patients, the C4-carnitine increase was not as strong even though significance was found. Furthermore, in both cases C4-carnitine and iso-C4-carnitine high levels were partially explained by age correction, indicating age related changes in energy metabolism. Finally, no significance was found in severe cases with poor outcome compared to discharged patients. This indicates that elevated levels are mainly associated with energy metabolism changes in severe disease occurrence.

#### 3.2.3 3’,4’-didehydro-3’-deoxycytidine (ddhC)

The last compound we selected to quantify in this study is a less well-known metabolite, ddhC, whose function and origin as a product of the viperin enzyme was elucidated recently. Viperin is part of radical *S*-adenosyl-l-methionine enzyme family and is known to be activated by interferon upon viral infection as part of a natural defence response (Rivera-Serrano et al., 2020). More recently, the mechanism by which viperin impedes viral proliferation was elucidated as being an enzymatic transformation of cytidine triphosphate (CTP) to 3’-deoxy-3’,4’-didehydro-cytidine triphosphate (ddhCTP) (Gizzi et al., 2018). The nucleotide version ddhC, has been shown in experimental studies to cross the cellular membrane and impede viral reproduction (Gizzi et al., 2018) even though the exact mechanism of this effect is contested. It was initially believed that ddhCTP served as a replication-chain terminator (Gizzi et al., 2018), however a recent study observed that ddhCTP is mostly inefficient as chain terminator and more likely impacts viral replication by depleting CTP and UDP pools in addition to impeding mitochondria function (Ebrahimi et al., 2020).

Elevated but non-quantitated serum levels of ddhC in COVID-19 patients have been previously reported by Mehta *et al*. (Mehta et al., 2022). Comparable to our results, ddhC showed exceptionally good capacity at distinguishing infected individuals from controls but its relation to disease severity was not so obvious. In the study performed by Mehta *et al*. (Mehta et al., 2022) ddhC was correlated with COVID-19 severity in the discovery cohort but not their validation cohort. In our data we found that the highest levels of ddhC in intermediate COVID-19 severity subgoup. These were patients that required respiratory support but not to the extent that patients with severe COVID-19; i.e., required FIO_2_ > 40% and/or required CPAP and/or required invasive ventilation. Moreover, when focusing our interest to severe infection cases only, poor outcome was associated with higher ddhC levels. This could indicate that ddhC is representative of viral infection level as directly triggered by viral presence, but not of the host ability to respond to that infection, where the perceived severity of the patient is a combination of viral proliferation and poorer host response.

Moreover, ddhC showed no correlation to any of the other compounds quantified in this study (SI Figure 19) or to any of the clinical data we considered (SI Figure 20). Unfortunately, no viral load data was available for our cohort to explore the likely correlation with ddhC. Those results indicate that ddhC could potentially hold mechanistic information in viral infection that is not currently represented by other measurements and as such it has some potential to support prognostic in combination with other markers in addition to a viral infection diagnostic.

#### 3.3 Limitations of the study

First, we would emphasise that infection severity labels are subjective and defined primarily by required level of oxygen support. In addition, these samples came from the first wave of COVID-19 and this lack of objectivity may in part be due to the surge in hospitalisation of patients and increased burden on critical care. Therefore, mild to intermediate to severe label change do not accurately present as a step change in biological processes. This is best illustrated by ddhC observed levels being highest in intermediate patients, but also KYN and KYN/TRP ratio progressive increase in intermediate cases. Unfortunately, this limitation in severity labels will negatively impact all regression models that explore the boundary between severe and non-severe infections.

In addition, a few more limitations of this study deserve to be discussed in more detail: a) the relatively small number of compounds that were selected for quantitation; b) the sample storage time; c) cohort size; and d) the reproducibility of the quantitative results in other laboratories.

In this study only a small number of compounds were selected to represent key pathways in COVID-19 infection and these were a subset of the compounds from the predictive model in our discovery study (Roberts et al., 2022). Those compounds offer a limited picture of complex host changes taking place during the infection. Moreover, metabolomics publications have also often referred to other compounds representing the amino acid metabolism and lipid markers (Costanzo et al., 2022; Mussap and Fanos, 2021) that deserve to be quantified and considered alongside this selection for clinical adoption.

The samples used in this study were the same as those used in the previous untargeted metabolomics study (Roberts et al., 2022). As validating a custom quantitative method required a significant time investment, sample storage time had to be extended. Even though samples were constantly kept at −80°C some compound degradation may be expected. This may have influenced the accuracy of the reported concentrations, but not the relative measurements as all samples were kept at the same conditions.

The study results are limited by the cohort size and more specifically by the small number of patients with poor outcome (*n*=23). This limitation is most visible when it comes to the exploration of multi-compound predictive models. Despite the interest of identifying the most predictive features between all proposed markers and available clinical measurements we kept the selection to a small number of handpicked features to avoid overturning to our data. We also purposefully did not use models requiring parameter tuning as our samples size would not permit a meaningful validation group, especially in outcome. However, we consider that our quantitative results could contribute to larger network studies aiming to explore such predictive modes further.

Finally, the quantitative results presented here were obtained using a custom LC-MS method. Even though the method went through an extensive validation process in our lab, it is important to reproduce those results on different cohorts by different labs and instruments.

## 4 Conclusions

This targeted, quantitative LC-MS study of control and COVID-19 patients serum samples suggests that KYNA/TRP ratio summarises best the changes that are taking place in TRP degradation pathway with severity and likelihood of poor outcome. Moreover, in multi-predictor models of outcome and severity KYNA/TRP showed the best individual performance and improved predictive performance of the models over clinical measurements alone.

With the exception of ddhC, the measured metabolites concentration in mild and discharged COVID-19 patients were comparable to control samples, indicating that C4-carnitines and TRP degradation upregulation takes place only in severe COVID-19 patients especially with higher chances for poor outcome. ddhC appears to be a promising indicator of a viral infection as levels in controls are nearly non-existent.

It is to be hoped that such accurate quantitative measurements for compounds frequently associated with COVID-19 disease development will allow those findings to move closer to medical practice adoption.

## 5 Materials and Methods

Internal labelled standards (ISTD), L-carnitine:HCl, O-butyryl (N-methyl-*d*_3_), cytidine (^15^N_3_), Kynurenic acid (Ring-*d*_5_), L-kynurenine sulfate (Ring-*d*_4_, 3,3-*d*_2_) were purchased by Cambridge Isotope Laboratories Inc. all certified at ≥ 98% chemical purity with the exception of kynurenine-*d*_6_ (≥ 95% purity). L-tryptophan - (indole-*d*5) (97% isotopic purity) was purchased from Sigma-Aldrich. Calibration curve standards for butyrylcarnitine, iso-butyrylcarnitine, cytidine, kynurenic acid, kynurenine and DL-Tryptophan were purchased from Sigma-Aldrich. 3’,4’-Didehydro-3’-deoxycytidine (ddhC) (CAS# 386264-46-6) was purchased from LGC standards GmbH. Commercial serum used as quality control in the study was pooled from BioIVT, Lot BRH1413770, Cat: HMSRM, mixed gender 0.1 µm filtered.

### 5.1 Sample acquisition and preparation for metabolomic analysis

COVID-19 patient and control serum samples were acquired as described in (Roberts et al., 2022). Ethical approval for the use of serum samples and associated metadata in this study was obtained from North West – Haydock research ethics committee (REC ref: 20/NW/0332). Control samples were confirmed by negative PCR test using Aptima SARS-CoV-2 assay on Hologic Panther platform. Sample preparation by protein precipitation in methanol was performed as described in (Roberts et al., 2022). For targeted analysis 40 µL of sample extract was spiked with an internal standard mixture to a final concentration of 0.063 µM butyrylcarnitine *d*_3_, 0.249 µM cytidine ^15^N_3_, 0.125 µM kynurenic acid *d*_5_, 2.5 µM kynurenine *d*_6_ and 50 µM tryptophan *d*_5_. The internal standard compound concentrations used were selected within the linear range of each compound and reflect the average concentration found in COVID-19 pooled samples during the method validation stage.

A 9-point calibration curve plus blank was prepared in PBS + 1% BSA (replacement matrix) in duplicates. The calibration curve started at 1 µM concentration for C4-carnitine, iso-C4-carnitine, cytidine, ddhC and KYNA. The highest concentration for KYN was 10 µM and 100 µM for TRP. The subsequent calibration points were a serial dilution as per the following example for 1 µM starting point, 0.8, 0.5, 0.4, 0.25, 0.125, 0.063, 0.031, 0.016, 0 µM. A detailed table per compound concentration can be found in the supplementary information SI Table 1. Standard addition QC samples were prepared in commercial serum and replacement matrix for all levels of the calibration curve points and then extracted by the same protein precipitation protocol.

Samples were analysed in four batches of 96 well plates, where each plate had two preparation replicates of the calibration curve and the QC samples (serum and replacement matrix), a plate pool QC, a study pool QC and 36 study samples. A plate pool QC was prepared by taking 5 µL from all patient samples on the plate while the study pool QC was obtained by mixing equal amounts of plate QCs. This was used to check reproducibility of quantitative results between plates. A template plate layout, run order and description of samples is provided in the <u>SI Table 2 and SI Appendix A respectively</u>. Samples, calibration curve mixtures and internal standards mixtures were all maintained on ice throughout the sample preparation. Complete blanks and internal standard spiked blanks were prepared in the same way replacing the matrix with water (LC–MS grade).

Plates were dried in a vacuum centrifuge (ScanVac MaxiVac Beta Vacuum Concentrator system, LaboGene ApS, Denmark) with no temperature application and stored at − 80 °C until required for UHPLC-MS/MS analysis. Prior to analysis, samples were resuspended in 40 µL water (LC–MS) for an injection volume of 2 µL, then centrifuged at 1,000×*g* for 5 min at 4 °C to remove air bubbles resulting from the resuspension.

### 5.2 UHPLC-MS/MS analysis of patient serum samples

Targeted UHPLC-MS/MS data acquisition was performed on a ThermoFisher Scientific Vanquish UHPLC system coupled to a ThermoFisher Scientific Q-Exactive mass spectrometer (ThermoFisher Scientific, UK). LC separation was performed in water (Solvent A) and methanol (solvent B) both with 0.1% formic acid on a Hypersil GOLD aQ C18 100×2.1mm, 1.9 um (ThermoFisher Scientific, UK) column at 50°C. A 10 min gradient with a flow rate of 0.4 mL/min was used. Gradient elution started at 1% B with an increase to 50% B between 2 to 6 min followed by a linear increase to 99% B from 6 at 6.5 min and held at this ratio to 8 min. Finally, the gradient was quickly brought back down to 1% B at 8.5 min and re-equilibrated at 1% B to the end of the method at 10 min.

The mass spectrometer source was set at C position with 3.2 kV spray voltage, 350 °C capillary temperature, 400 °C aux gas heater temperature, 48, 15 and 0 arbitrary unit for sheath, aux and sweep gas flow respectively. Finally, S-lens RF was set to 60%. A parallel reaction monitoring (PRM) mass spectrometry method was used at 17,500 FWHM at m/z 200 orbitrap resolution and 1.2 *m/z* quadrupole isolation window. The list of targeted compounds, their expected elution time and the collision energies used can be found in Table 5.

**Table 5.** MS method details per compound including retention time (RT), collision energy (N)CE in higher energy C trap dissociation (HCD) cell and parallel reaction monitoring (PRM) as precursor, quantitative target, confirming ion and fragment when applicable.

| Compound | Formula | Polarity | Start time (min) | End time (min) | (N)CE (V) | Precursor ($m/z$) | Target ($m/z$) | Confirming ($m/z$) | Fragment ($m/z$) |
| --- | --- | --- | --- | --- | --- | --- | --- | --- | --- |
| Butyrylcarnitine | $C_{11}H_{21}NO_4$ | ESI+ | 3.50 | 4.30 | 30 | 232.154 | 85.028 | 173.081 | 60.081 |
| Iso-butyrylcarnitine | $C_{11}H_{21}NO_4$ | ESI+ | 3.50 | 4.30 | 30 | 232.154 | 85.028 | 173.081 | |
| Butyrylcarnitine $d_3$ | $C_{11}H_{18}D_3NO_4$ | ESI+ | 3.50 | 4.30 | 30 | 235.173 | 85.030 | 173.130 | |
| Cytidine | $C_9H_{13}N_3O_5$ | ESI+ | 0.40 | 1.50 | 90 | 244.090 | 112.050 | 95.024 | 69.450 |
| Cytidine $^{15}N_3$ | $C_9H_{13}^{15}N_3O_5$ | ESI+ | 0.40 | 1.50 | 90 | 247.080 | 115.042 | 97.018 | 71.039 |
| ddhC | $C_9H_{11}N_3O_4$ | ESI+ | 0.60 | 1.50 | 30 | 226.082 | 112.050 | 190.061 | 147.055 |
| Kynurenic acid | $C_{10}H_7NO_3$ | ESI+ | 4.70 | 5.50 | 80 | 190.050 | 162.055 | 116.049 | 89.038 |
| Kynurenic acid $d_5$ | $C_{10}H_2D_5NO_3$ | ESI+ | 4.70 | 5.50 | 80 | 195.081 | 167.086 | 121.081 | 94.070 |
| Kynurenine | $C_{10}H_{12}N_2O_3$ | ESI+ | 1.50 | 3.50 | 30 | 209.092 | 94.065 | 192.065 | 146.060 |
| Kynurenine $d_6$ | $C_{10}H_6D_6N_2O_3$ | ESI+ | 1.50 | 3.50 | 30 | 215.130 | 98.090 | 198.103 | |
| Tryptophan | $C_{11}H_{12}N_2O_2$ | ESI+ | 4.10 | 4.70 | 30 | 205.097 | 188.070 | 146.060 | 118.065 |
| Tryptophan $d_5$ | $C_{11}H_7D_5N_2O_2$ | ESI+ | 4.10 | 4.70 | 30 | 210.129 | 192.095 | 150.085 | 122.090 |

Calibration curves and spiked replacement matrix QCs were run in three technical replicates. Two replicates were acquired at the beginning of the batch and one at the end. Similarly, two replicate injections of commercial serum spiked QCs were acquired at the beginning of the batch and at the end. Thirty-six patient samples were loaded per plate of which 6 randomly selected samples were injected in duplicate and 30 in singlets for a total plate runtime of 39 hours. This approach of restricting sample replication was taken because the method validation tests showed excellent reproducibility between injections and allowed us to minimize time-based drift effects in the study.

### 5.3 Data processing

Raw data were processed in Thermo Fisher scientific Trace Finder (Version 5.1 Build 203). Compound detection was performed based on the target and confirming ion transitions listed in Table 5. Compound concentrations were normalized based on internal standards, with iso-C4-carnitine normalized on butyrylcarnitine-*d*3 and ddhC normalized on cytidine ^15^N_3_ due to difficulty of obtaining a labelled version of those compounds.

Matrix factor corrections were calculated and applied to all study samples based on the peak area differences in spiked commercial serum samples *vs*. replacement matrix. The peak area corresponding to the addition of the standard is calculated by subtracting the peak area of non-spiked commercial serum sample. Normalized matrix factor, calculated by dividing the peak ratio of the standard over the peak ratio of the labelled standard was used for correction. Finally, an average of the normalized ratios was calculated over multiple spike levels. The spike levels used to calculate the average per compound were defined based on lower limit of quantification (LLoQ) and upper limit of quantification (ULoQ) of that compound.

Data quality was assessed based on calibration curve linearity, precision of technical replicates and accuracy of spiked sample readings following FDA guidelines described in (FDA, 2018). Finally relative standard deviation (RSD) between plates of study pooled QC and commercial serum QC were used to assess quantitative precision between batches.

### 5.4 Statistical data analysis

Statistical analysis was performed in the same manner as the preceding untargeted discovery study (Roberts et al., 2022). All the packages, versions and code were the same and are available in github (https://github.com/dbkgroup/COVID). In brief, the following approach was adopted: individual odds ratios (OR) and 90% confidence intervals (CI) for selected compounds were determined using Bayesian logistic regression from the *stan_glm* function in the *rstanarm* R package (Gabry and Goodrich, 2020). Compounds were adjusted for age, sex and BMI. Results for multiple compound models were produced with a simple additive Bayesian logistic regression model using the same package. Data were autoscaled to have mean = 0 and standard deviation (SD) = 1 to allow comparison of ORs. When present, replicated samples were averaged prior to modelling. To avoid overfitting (Broadhurst and Kell, 2006) and evaluate the model sensitivity to the data, Monte Carlo cross validation, with 100 iterations and a 70:30 train test split, was used and prediction metrics were reported as mean and SD. Conservative regularization parameters were used to reduce overfitting with prior scale = 1. No model tuning or hyperparameter optimization was applied.

## Data Availability

UHPLC-MS data for the quantitative study in RAW format is freely available in the MetaboLights repository (Haug et al., 2020) with study identifier MTBLS7429 (https://www.ebi.ac.uk/metabolights/).

## Author Contributions

D.B.K. and R.G. conceived the study and obtained funding for a grant to enable this work.

A.S.D and J.M.T. obtained ethical approval, acquired samples and metadata. I.R., M.W.M. and C.L.W designed targeted quantitative UHPLC-MS/MS method.

I.R. and M.W.M prepared samples and performed UHPLC-MS/MS acquisition.

I.R. processed data and performed data analysis.

I.R. performed data interpretation and wrote the manuscript with M.W.M and D.B.K. All authors read and approved the final version of the manuscript.

## Ethical Statements

All procedures performed in studies involving human participants were in accordance with the ethical standards of the institutional and/or national research committee and with the 1964 Helsinki declaration and its later amendments or comparable ethical standards.

Ethical approval for the use of serum samples and associated metadata in this study was obtained from North West – Haydock research ethics committee (REC ref: 20/NW/0332).

