## Supplementary information for "Quantitative LC-MS study of compounds found predictive of COVID-19 severity and outcome"

Ivayla Roberts, Marina Wright Muelas, Joseph M. Taylor, Andrew S. Davison, Catherine L. Winder, Royston Goodacre & Douglas B. Kell

#### 1 Supplementary background

##### 1.1 3',4'-didehydro-3'-deoxycytidine (ddhC) identification

In the untargeted discovery LC-MS study we retained the use of an in-source cytosine fragment in the model [1]. At that time, we believed that this fragment was produced by deoxycytidine as it fit well based on retention tine (RT) and a strong tendency to fragment at the source. However, further investigation while preparing the quantitative method showed that the fragment is in fact ddhC a deoxycytidine analogue. The identification method followed here was similar but independent to the identification described in [2]. This independent identification increases the likelihood of the conclusion to be correct and confirms the importance of ddhC in viral infections.

When it comes to COVID-19, cytosine has been reported as relevant in a few metabolomics studies. Indeed, multiple cytosine events can be observed in the patient's serum in our study. However, it became obvious that only one of those events could be attributed to cytosine and others are more likely a source fragmentation of cytosine substructures of more complex molecules (see SI Figure 1). After comparison against standards, we established the most likely origin of some of the fragments.

The two signals shown in SI Figure 1 result from QC (COVID-19 patients serum pool) and SQA (commercial serum quality control) samples. We can see the 4 events of 112.0505 *m/z* (cytosine) are increased in COVID-19 patients at RT of 0.758, 0.840, 0.920, 1.317, 1.396 min. All those compounds eluted early and are hardly influenced by the column but remained well grouped and reproducible between the 4 batches of samples composing the study in question. As a reminder the column used in the study is Hypersil GOLD aQ C18 (ThermoFisher Scientific, UK) with a gradient at 1%B (MeOH + 0.1% formic acid) at the time of elution of all cytosine events.

The events at RT 0.840 & 0.920 min were both retained as good quality peaks and identified in Compound Discoverer (ThermoFisher Scientific version 3.1) as cytosine against mzCloud with a score of 73.9%; (good match). These events also scored 76.7% in spectral similarity against a cytosine standard (Sigma Aldrich) run on the same MS instrument. The events at 0.728, 1.317 and 1.396 min were not identified as good quality peaks and therefore discarded. Their intensity did not allow for MS2 spectra acquisition.

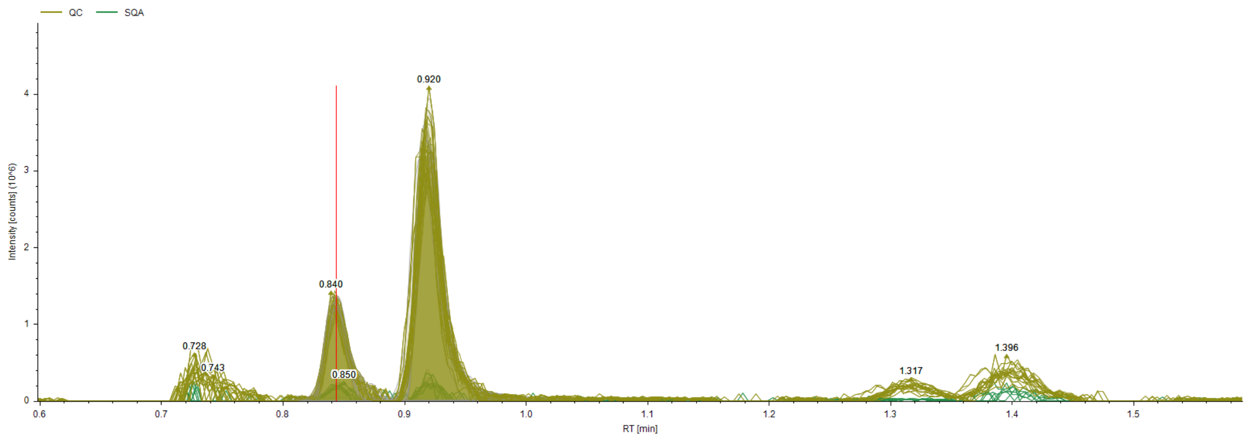


*SI Figure 1. 112.0505 m/z events (cytosine like) in COVID-19 patients pooled sample.*

Furthermore, a comparison of RT for cytosine, cytidine and deoxycytidine showed that cytidine is the first to elute at ~0.7 min, followed by cytidine and deoxycytidine. This is demonstrated in SI Figure 2 with pooled COVID-19 patients' serum (in black) and same serum spiked with 5 µM of cytidine, cytosine and deoxycytidine mix (in red). Both signals are filtered for mass range 112.05 – 112.051 (cytosine *m/z* is of 112.0505). The standards were also run separately allowing to establish the RT for each. We can also observe that for the same concentration levels deoxycytidine produces the strongest signal of cytosine fragment. Indicating that those are the most likely origins.

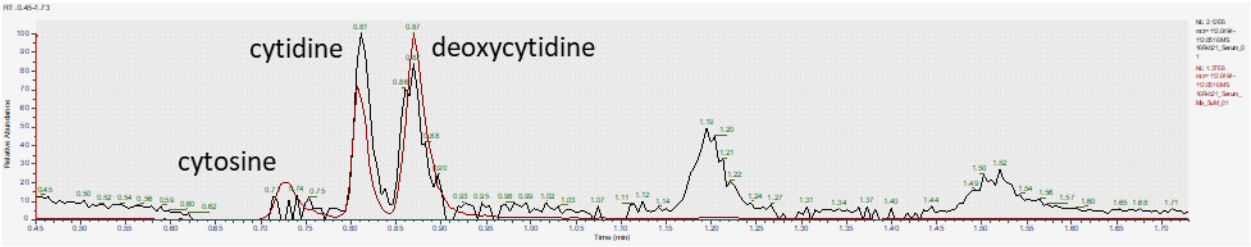


*SI Figure 2. COVID-19 patients pooled serum (black) and COVID-19 patients pooled serum spiked with cytosine, cytidine and deoxycytidine.*

However, further investigation showed that the cytidine fragment of interest was most likely resulting from an unknown compound with calculated mass of 225.0747 and predicted composition of $C_9H_{11}N_3O_4$. The unknown compound also showed cytosine fragment in its MS/MS spectra. Moreover, the ion counts of the compound showed a $R^2$ of 0.93 (SI Figure 3). After a more detailed look into the compound fragmentation, it showed 2 water loss events indicating high likelihood of 2 OH groups and predicted composition similar to dCytdine minus 2H. An exploration of structural similarity and predicted fragmentation indicated ddhC a good candidate (SI Figure 4).

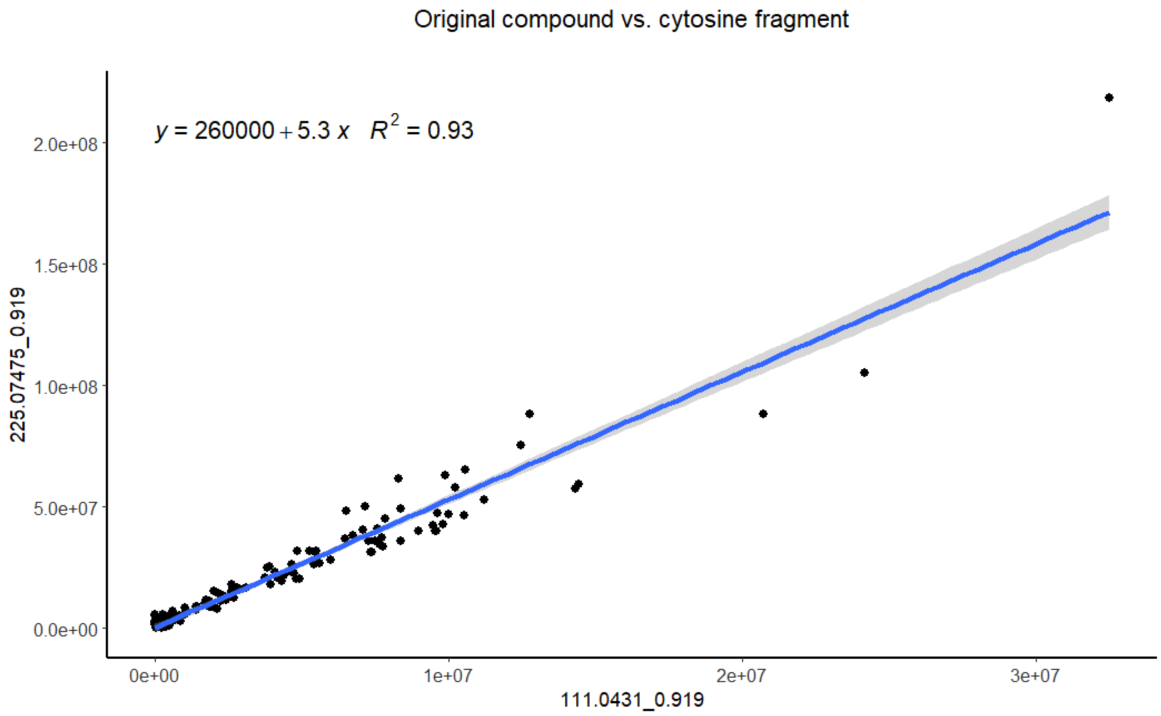


*SI Figure 3. Normalized peak areas of the fragment of interest 111.0431 m/z @ 0.919 min RT and the most likely source – unknown compound of 225.0775 m/z @ 0.919 min RT. The area corelation shows an $R^2$ of 0.93*

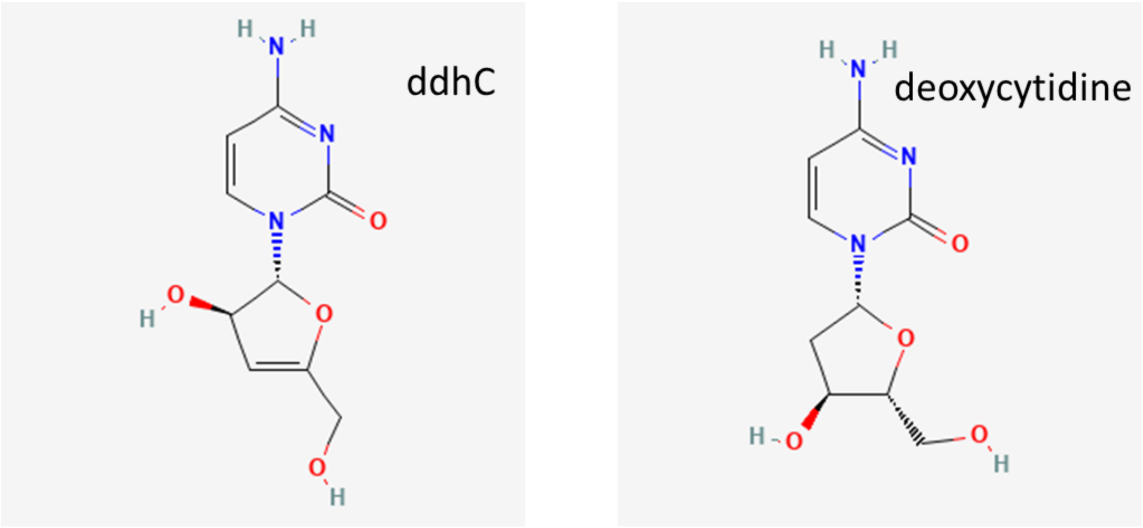


*SI Figure 4. Side-by-side comparison of the structure of ddhC and deoxycytidine*

Following standard acquisition and extensive confirmation of similarity of fragmentation at different collision energies, the identification was confirmed as ddhC and the compound was included in the quantitative study.

DdHC MS/MS spectra obtained by varying the energy in higher-energy collisional dissociation (HCD) C-trap on a ThermoFisher Scientific ID-X Tribrid mass spectrometer (ThermoFisher Scientific, UK) is presented in SI Figure 5. With (a) HCD 10, (b) HCD 40, (c) HCD 60 and (d) HCD 80.

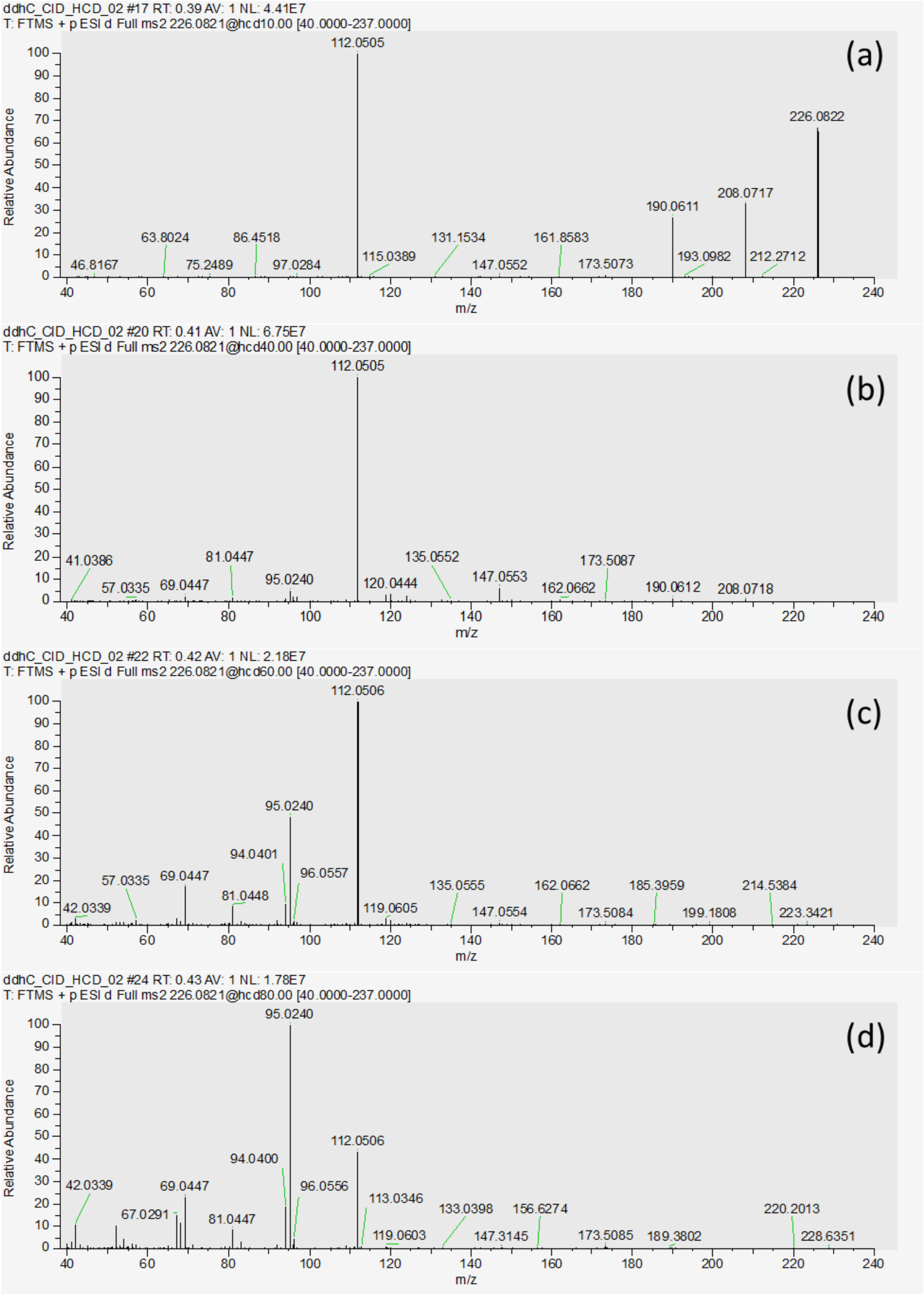


*SI Figure 5. ddhC standards MS/MS spectrum at (a) HCD 10, (b) HCD 40, (c) HCD 60 and (d) HCD 80.*

As shown in our results, true cytosine is hardly detected in our COVID-19 study. However, cytosine source fragments from ddhC, and most likely cytidine (identity not confirmed), are detected. Following these results, we would recommend the community to carefully verify future cytosine findings in infectious disease.

### 2 Supplementary methods

*SI Table 1. Calibration curve concentrations per compound in µM*

| Compound | C1 (µM) | C2 (µM) : (80% C1) | C3 (µM) (50% C1) | C4 (µM) (50% C2) | C5 (µM) (50% C3) | C6 (µM) (50% C5) | C7 (µM) (50% C6) | C8 (µM) (50% C7) | C9 (µM) (50% C8) | C0 (µM) |
|---|---|---|---|---|---|---|---|---|---|---|
| **Butyrilcarntine** | 1 | 0.8 | 0.5 | 0.4 | 0.25 | 0.125 | 0.063 | 0.031 | 0.016 | 0 |
| **Iso-Butyrylcarnitine** | 1 | 0.8 | 0.5 | 0.4 | 0.25 | 0.125 | 0.063 | 0.031 | 0.016 | 0 |
| **Cytidine** | 1 | 0.8 | 0.5 | 0.4 | 0.25 | 0.125 | 0.063 | 0.031 | 0.016 | 0 |
| **ddhC** | 1 | 0.8 | 0.5 | 0.4 | 0.25 | 0.125 | 0.063 | 0.031 | 0.016 | 0 |
| **KA** | 1 | 0.8 | 0.5 | 0.4 | 0.25 | 0.125 | 0.063 | 0.031 | 0.016 | 0 |
| **Kynurenine** | 10 | 8 | 5 | 4 | 2.5 | 1.25 | 0.63 | 0.31 | 0.16 | 0 |
| **Tryptophan** | 100 | 80 | 50 | 40 | 25 | 12.5 | 6.3 | 3.1 | 1.6 | 0 |

*SI Table 2. Plate layout template*

| Row / Col | 1 | 2 | 3 | 4 | 5 | 6 | 7 | 8 | 9 | 10 | 11 | 12 |
|---|---|---|---|---|---|---|---|---|---|---|---|---|
| A | BC0 | BC9 | BC8 | BC7 | BC6 | BC5 | BC4 | BC3 | BC2 | BC1 | Blank_ISTD | Blank |
| B | BCQC | BCQC | BCQ_none | BQC_C9 | BQC_C8 | BQC_C7 | BQC_C6 | BQC_C5 | BQC_C4 | BQC_C3 | BQC_C2 | BQC_C1 |
| C | SCQC | SQC_none | SQC_C9 | SQC_C8 | SQC_C7 | SQC_C6 | SQC_C5 | SQC_C4 | SQC_C3 | SQC_C2 | SQC_C1 | QC_study |
| D | S1 | S2 | S3 | S4 | S5 | S6 | S7 | S8 | S9 | S10 | S11 | S12 |
| E | S13 | S14 | S15 | S16 | S17 | S18 | S19 | S20 | S21 | S22 | S23 | S24 |
| F | S25 | S26 | S27 | S28 | S29 | S30 | S31 | S32 | S33 | S34 | S35 | S36 |
| G | BC0 | BC9 | BC8 | BC7 | BC6 | BC5 | BC4 | BC3 | BC2 | BC1 | QC_plate | QC_plate |
| H | BQC_C9 | BQC_C8 | BQC_C7 | BQC_C5 | BQC_C3 | BQC_C2 | SQC_C9 | SQC_C8 | SQC_C7 | SQC_C5 | SQC_C3 | SQC_C2 |

| | |
|---|---|
| BC# | PBS + 1%BSA replacement matrix calibration curve point |
| BCQC | PBS + 1%BSA replacement matrix conditioning sample |
| BQC_C# | PBS + 1%BSA replacement matrix spiked at calibration point level QC |
| SCQC | Commercial serum conditioning sample |
| SQC_C# | Commercial serum QC spiked at calibration point level |
| QC_plate | Pool of plate study samples |
| QC_study | Pool of all study samples |
| S## | Study sample |

### 3 Supplementary results

This section contains supplementary information on the targeted quantitative LC-MS method followed by additional figures and tables supporting the statistical data analysis.

#### 3.1 Quantitative data quality

The targeted quantitative LC-MS method was developed to allow accurate measurement of 6 compounds, kynurenic acid (KYNA), kynurenine (KYN), tryptophan (TRP), butyrylcarnitine (C4-carnitine), iso-butyrylcarnitine (iso-C4-carnitine) and 3',4'-didehydro-3'-deoxycytidine (ddhC). The method was validated at 5 different occasions ahead of the study and within the study based on calibration curves linearity, accuracy, precision, and reproducibility. The data quality assessment within the study is presented in this section.

Compound elution times (SI Figure 6) were well spread over the 10 min LC gradient. ddhC elute early and were barely retained by the column but remained reproducible. C4-carnitiens elutes closely after its isomeric form, however, remains well separated in retention time (RT) as was also reported by [3]. KYNA showed peak tailing tendency in this method; however, its reproducibility and linearity were impeccable; therefore, this was judged satisfactory for the purpose of this study.

The sample dilution level was selected to accommodate most of the compounds which excluded cytidine despite our attempt to add it to the method. No common sample dilution level could offer tryptophan and cytidine linearity at the same time. This could be seen in the calibration curve linearity drop and increased QC RSDs for cytidine (SI Figure 12 and SI Table 5). As results cytidine was dropped from the data analysis. ddhC quantitation was also negatively impacted by the sample dilution levels, however, ddhC data quality attributes are still acceptable therefore we have chosen to report the results with a warning on the higher uncertainty surrounding the concentration for this compound.

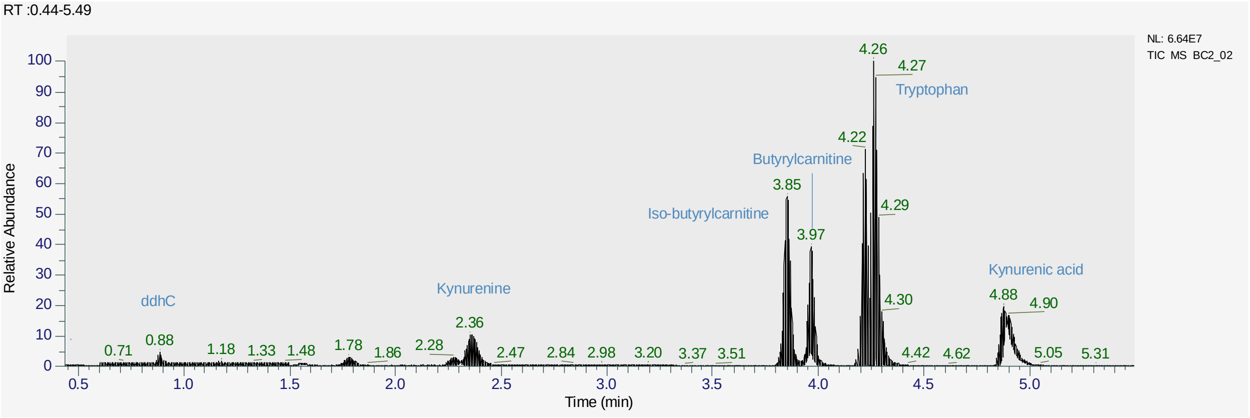


*SI Figure 6. Total Ion Chromatogram (TIC) and Retention Time (RT) for the targeted compounds in the method.*

<u>Calibration curves</u>

Most of the compounds showed good calibration curve linearity with $R^2$ values > 0.99 per plate and all plates together (SI Figure 7 – SI Figure 13), except cytidine and ddhC where the $R^2$ dropped to 0.98 in average as showed in SI Figure 12 and SI Figure 13. The drop in performance for cytidine and ddhC appears to be mostly due to the suboptimal performance of cytidine and cytidine [15]N3 in this method at the chosen sample concentration level. ddhC performance of non-normalized peak areas shows a $R^2$ > 0.99 in three plates out of the four plates and > 0.98 in the remaining batch.

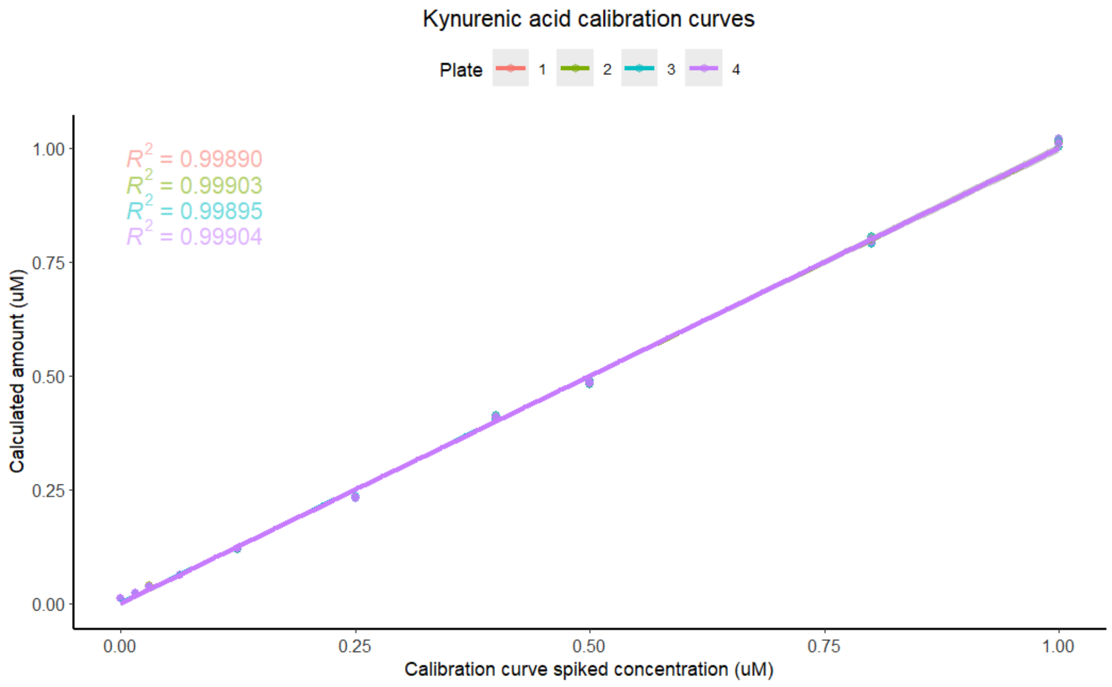


*SI Figure 7. KYNA calibration curves per study plate*

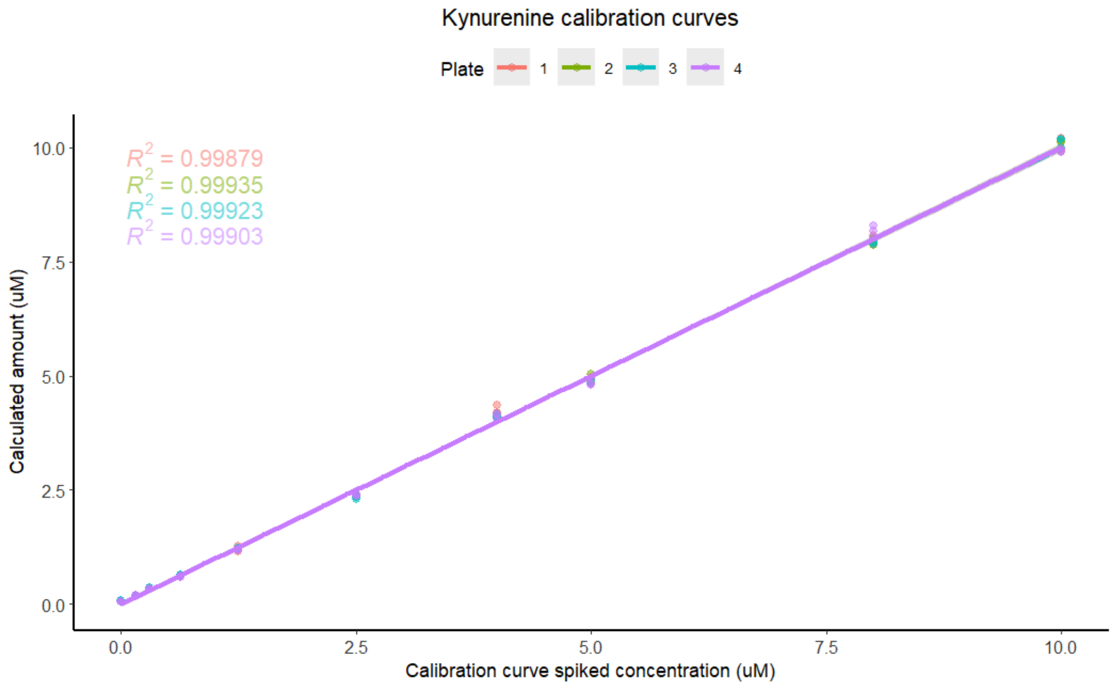


*SI Figure 8. KYN calibration curves per study plate*

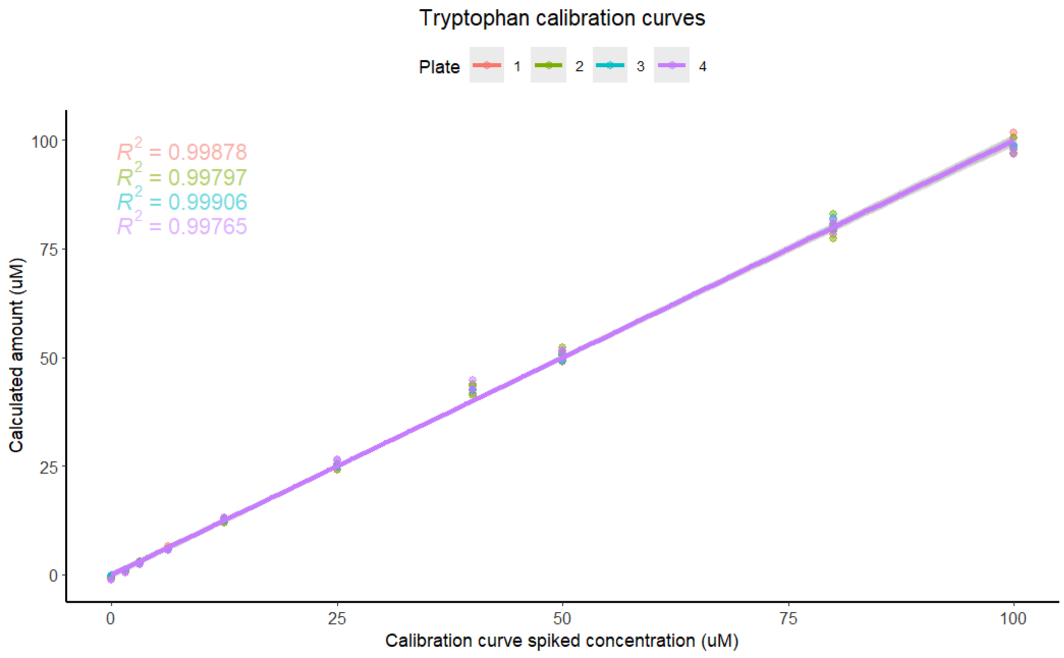


*SI Figure 9. TRP calibration curves per study plate.*

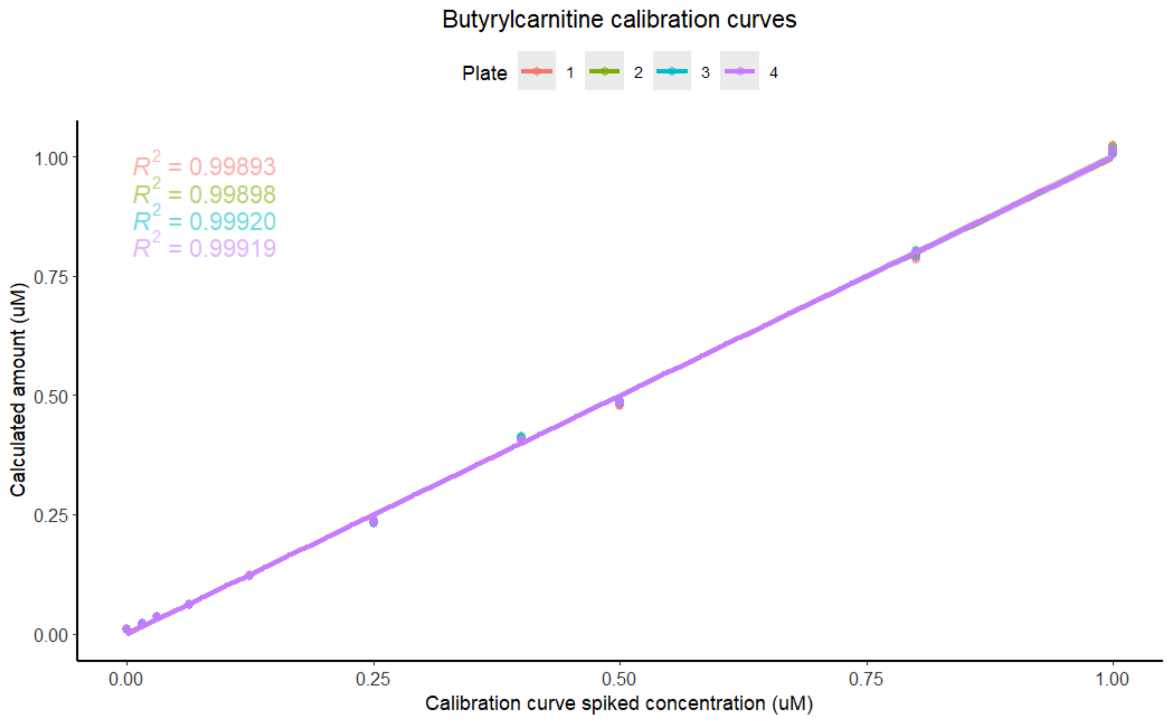


*SI Figure 10. C4-carnitine calibration curves per study plate*

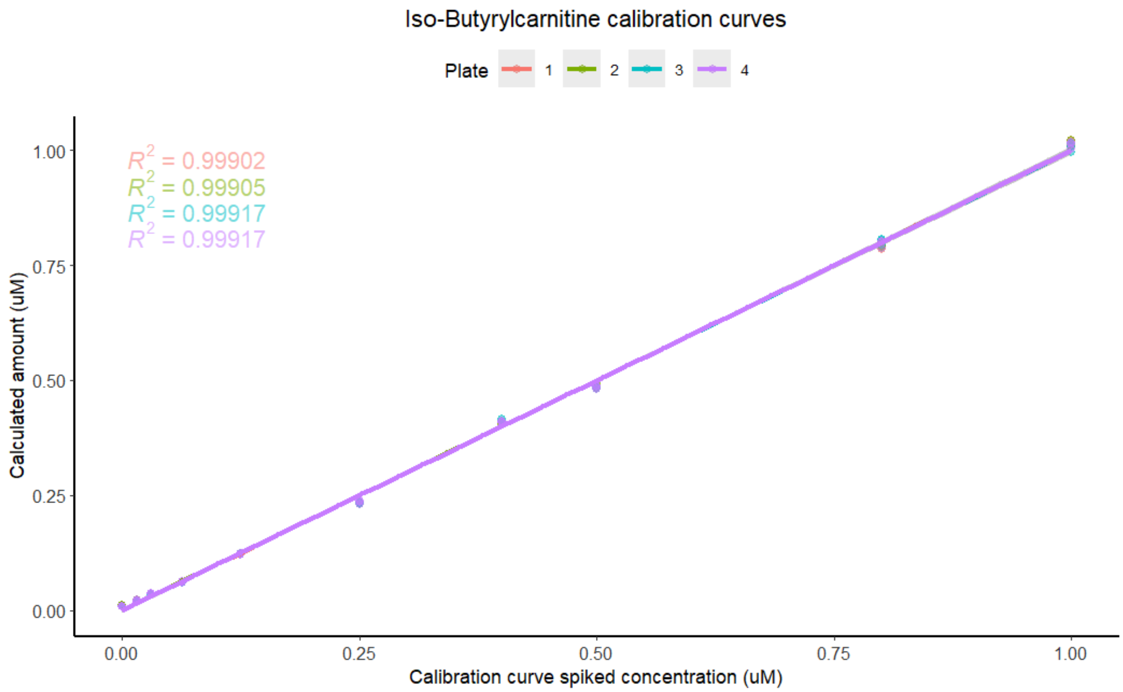


*SI Figure 11. Iso-C4-carnitine calibration curves per study plate*

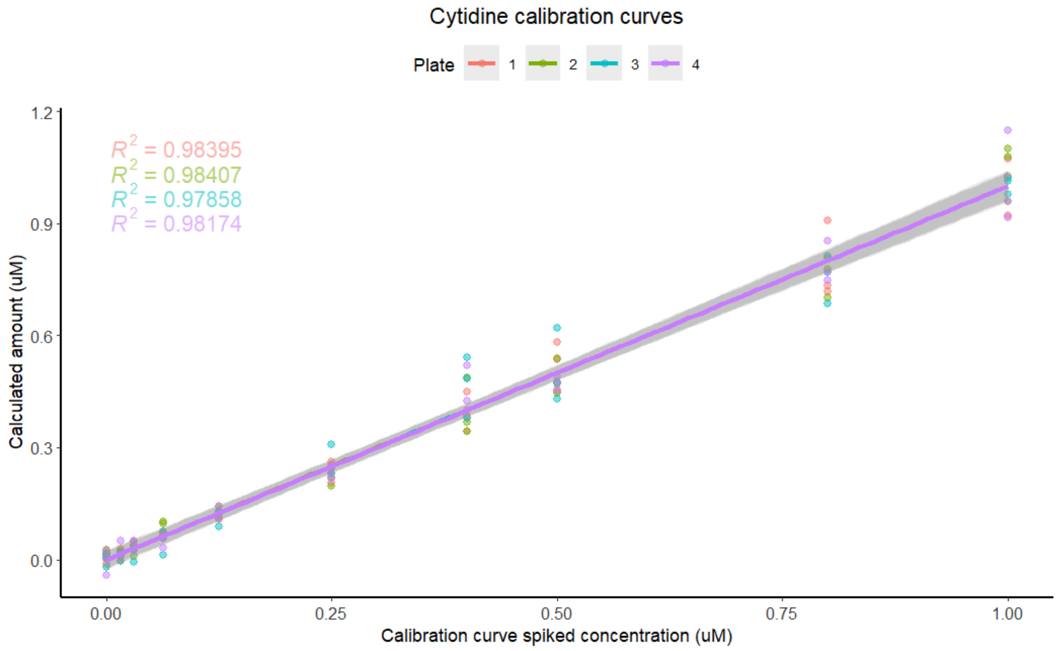


*SI Figure 7. Cytidine calibration curves per study plate*

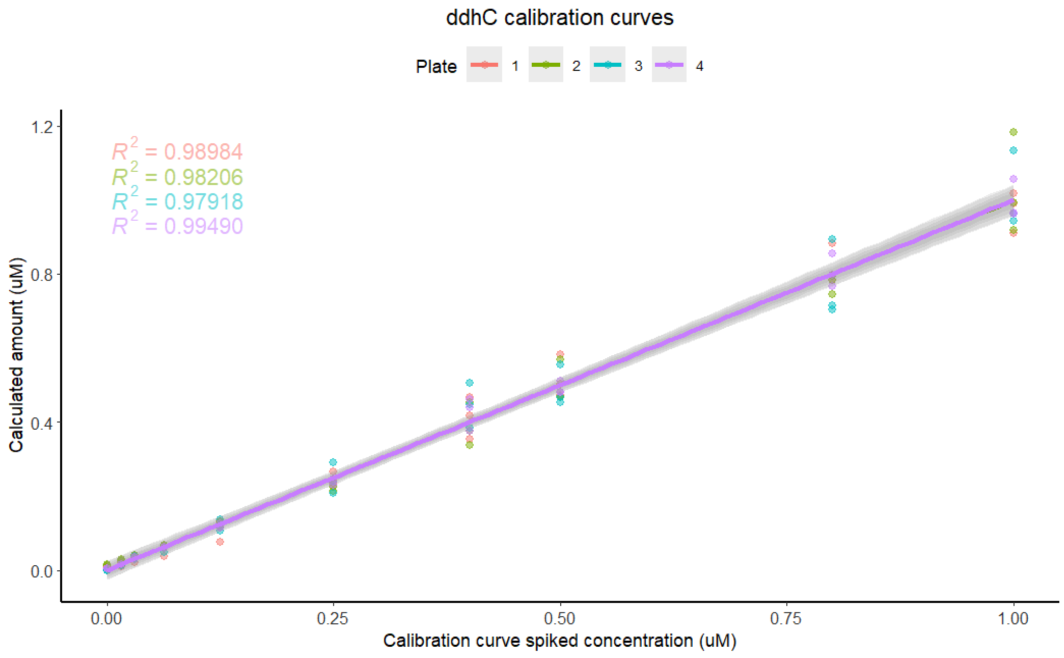


*SI Figure 8. ddhC calibration curves per study plate*

#### Matrix factor

Matrix factor (MF) correction was performed for all calculated compound concentrations in serum as described in the main paper methods section. SI Table 3 shows the calculated MF per plate for each compound and the spiked QC levels used in the presented average. Low level calibration points were removed due to poor accuracy. For most compounds C1 to C7 spike levels were retained with noticeable exception in the case of cytidine where C1-C4 levels only offered an acceptable accuracy. The MF was close to 1 for most of the compounds at the exception of KYN where MF was consistently close to 3-fold difference.

*SI Table 3 Matrix factor correction per target compound*

| Compound | Spike levels used | Plate 1 | Plate 2 | Plate 3 | Plate 4 |
|---|---|---|---|---|---|
| **KYNA** | C1-C7 | 1.01 | 1.00 | 0.97 | 1.00 |
| **KYN** | C1-C6 | 2.93 | 2.93 | 2.85 | 2.86 |
| **TRP** | C1-C6 | 0.89 | 0.77 | 0.81 | 0.85 |
| **C4-carnitine** | C1-C7 | 0.82 | 0.88 | 0.95 | 0.96 |
| **iso-C4-carnitine** | C1-C7 | 0.83 | 0.90 | 0.96 | 0.98 |
| **ddhC** | C1-C7 | 0.88 | 0.97 | 0.78 | 1.14 |
| **cytidine** | C1-C4 | 0.98 | 0.90 | 0.98 | 0.92 |

Accuracy

Accuracy was calculated based on compound concentration results in commercial serum QC samples spiked with known amount of compound standard. The predicted standard addition was obtained by subtracting the endogenous concentration calculated in non-spiked QC sample from the spiked QC sample. Accuracy of quantitation was calculated as % difference between estimated standard addition versus the known standard amount addition. A difference lower than 15% is desired where 20% difference is tolerated at the lower limit of quantitation (LLOQ) level.

As shown in SI Table 4, the lowest level of accurate quantitation of TRP was at 12 µM (C6 level), C4, iso-C4 carnitine, and ddhC LLOQ was reached between 0.031 and 0.016 µM (C8-C9). KYNA crossed LLOQ between 0.06 – 0.031 µM (C7-C8) and KYN accuracy remained in good down to 0.31 µM (C8). Finally, cytidine shows steep accuracy drop between 0.125 – 0.063 µM, so as high % difference at concentrations above 0.8 µM. This supports the conclusion that the sample dilution levels are unsuitable for cytidine quantitation.

*SI Table 4 Accuracy metrics per compounds in serum QC samples*

| Compound | QC C9 Diff (%) | QC C8 Diff (%) | QC C7 Diff (%) | QC C6 Diff (%) | QC C5 Diff (%) | QC C4 Diff (%) | QC C3 Diff (%) | QC C2 Diff (%) | QC C1 Diff (%) |
|---|---|---|---|---|---|---|---|---|---|
| **KYNA** | 28.35 | 21.97 | 17.81 | 11.72 | 7.13 | 0.74 | 8.49 | 4.13 | 1.65 |
| **KYN** | 24.81 | 15.93 | 8.77 | 7.51 | 5.84 | 2.67 | 8.75 | 2.75 | 2.94 |
| **TRP** | 129.20 | 46.38 | 36.31 | 18.86 | 5.24 | 6.25 | 8.15 | 9.23 | 7.02 |
| **C4-carnitine** | 20.10 | 18.94 | 10.89 | 10.02 | 5.00 | 2.72 | 7.30 | 4.66 | 2.54 |
| **iso-C4-carnitine** | 23.63 | 18.30 | 10.72 | 9.43 | 4.65 | 3.12 | 7.22 | 4.56 | 2.93 |
| **ddhC** | 42.25 | 17.06 | 8.97 | 9.48 | 1.79 | 4.45 | 5.58 | 8.87 | 3.55 |
| **Cytidine** | NA | 48.63 | 32.69 | 6.19 | 3.32 | 12.00 | 4.42 | 16.63 | 14.31 |

Precision

Precision was calculated based the three replicate injections of commercial serum QC. Relative Standard Deviation (RSD) lower than 15% was desired, where 20% RSD was acceptable at LLOQ. For most of the compounds the RSD was exceptionally good (< 3%) as shown in SI Table 5. ddhC RSDs were consistently < 10% and cytidine RSDs were acceptable only for concentrations higher than 0.125 µM (C6) of added standard.

*SI Table 5. Precision metrics per compound in serum QC samples*

| Compound | QC C9 RSD (%) | QC C8 RSD (%) | QC C7 RSD (%) | QC C6 RSD (%) | QC C5 RSD (%) | QC C4 RSD (%) | QC C3 RSD (%) | QC C2 RSD (%) | QC C1 RSD (%) |
|---|---|---|---|---|---|---|---|---|---|
| **KYNA** | 0.33 | 0.54 | 0.94 | 0.7 | 0.72 | 0.68 | 0.29 | 0.88 | 0.72 |
| **KYN** | 1.03 | 0.6 | 1.12 | 1.41 | 2.15 | 1.54 | 1.22 | 0.94 | 1.32 |
| **TRP** | 0.84 | 0.99 | 1.19 | 0.63 | 0.64 | 1.6 | 1.22 | 1.66 | 1.36 |
| **C4-carnitine** | 0.23 | 2.46 | 0.78 | 2.75 | 0.47 | 4.9 | 0.37 | 5.01 | 1.01 |
| **iso-C4-carnitine** | 0.56 | 3.07 | 0.54 | 2.33 | 0.92 | 4.3 | 0.78 | 5.01 | 0.53 |
| **ddhC** | 5.74 | 8.33 | 7.73 | 6.58 | 4.88 | 5.4 | 4.64 | 9.38 | 8.31 |
| **Cytidine** | NA | -77.15 | -5 | 14.79 | 13.92 | 7.88 | 6.54 | 7.06 | 4.93 |

Quantitation reproducibility between plates (RSDs)

Finally, the quantitation reproducibility between plates was estimated based on commercial serum QC samples and study serum pooled QC. The RSDs between plates of the quantitative results is shown in SI Table 6; all compounds except cytidine have acceptable RSD <15%. The method did not detect cytidine in commercial serum.

*SI Table 6. Mean quantitation values and RSD in the study QCs*

| Compound | Study QC mean (μM) | Study QC RSD (%) | Commercial serum mean (μM) | Commercial serum RSD (%) |
|---|---|---|---|---|
| **KYNA** | 0.11 | 4.44 | 0.04 | 0 |
| **KYN** | 3.23 | 10.91 | 1.56 | 2.67 |
| **TRP** | 53.06 | 7.41 | 91.12 | 5.96 |
| **C4-carnitine** | 0.27 | 4.87 | 0.07 | 0 |
| **iso-C4-carnitine** | 0.18 | 4.54 | 0.09 | 5.71 |
| **ddhC** | 0.49 | 11.90 | 0.07 | 11.66 |
| **Cytidine** | 0.07 | 20.20 | - | - |

In conclusion, the quality of all compound quantitation data, except cytidine, showed satisfactory results. Therefore, cytidine was excluded from further analysis given its poor performance in precision, accuracy, and quantitation reproducibility in low concentrations i.e., < 0.125 uM which are of biological relevance as several cytidine measurement in the study were < 0.125 uM. All compounds showed measurements comparable with previously reported in the literature concentrations in commercial serum[4], except in the case of ddhC where no quantitative information was found.

#### 3.2 Statistical analysis

This section contains figure and tables supporting the statistical analysis section in the main paper.

##### 3.2.1 Mild and discharged COVID-19 patient's *vs.* control cases

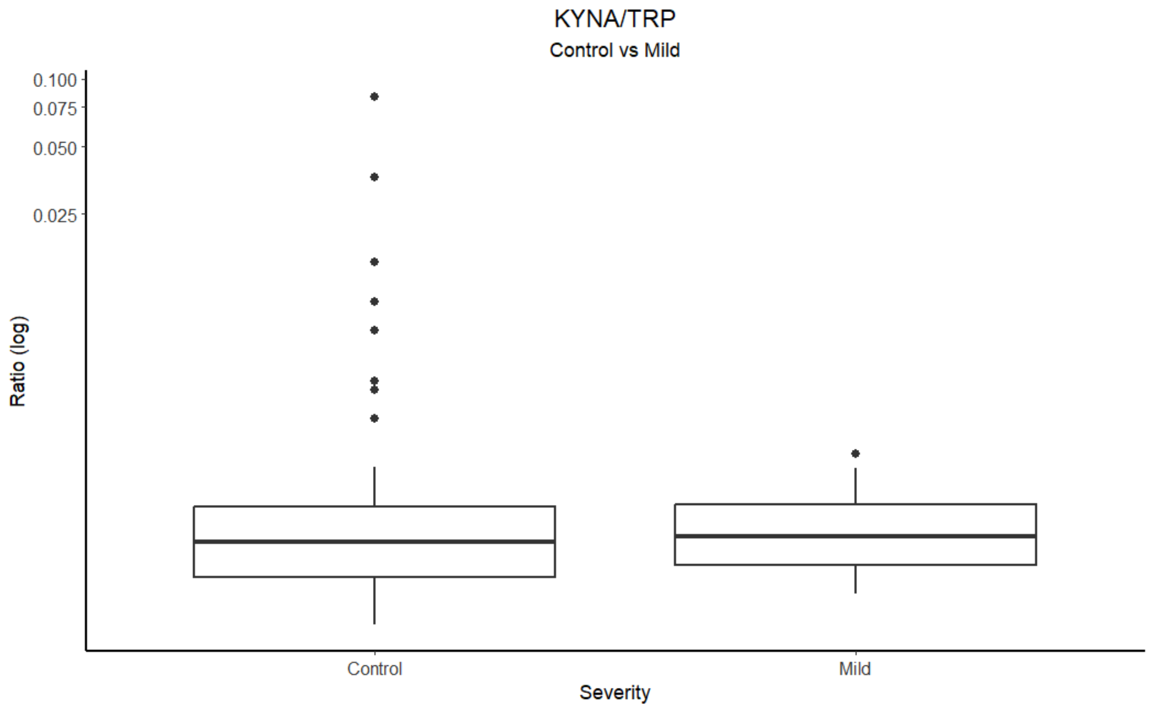


*SI Figure 9. KYNA/TRP ratio in log scale for control patient's vs mild COVID-19 cases. The group differences appear to be mainly driven by outlier in the control population.*

*SI Table 7. Logistic regression results corrected for age, sex and BMI in control patients compared to all COVID-19 patients, to mild COVIC-19 patients and discharged COVID-19 patients.*

| | Control vs Disease | | | | | Control vs Mild | | | | | Control vs Discharged | | | | |
|---|---|---|---|---|---|---|---|---|---|---|---|---|---|---|---|
| | **No correction** | **Age** | **Sex** | **BMI** | **Age, Sex & BMI** | **No correction** | **Age** | **Sex** | **BMI** | **Age, Sex & BMI** | **No correction** | **Age** | **Sex** | **BMI** | **Age, Sex & BMI** |
| **KYNA** | 0.85 (0.61-1.1) | 0.86 (0.62-1.2) | 0.83 (0.61-1.1) | 0.79 (0.54-1.1) | 0.76 (0.5-1.1) | 0.16 (0.017-0.64) | 0.21 (0.024-0.7) | 0.13 (0.013- | 0.21 (0.025- | 0.24 (0.029-0.76) | 0.65 (0.37-0.97) | 0.67 (0.4-1) | 0.63 (0.37-0.94) | 0.59 (0.33-0.93) | 0.57 (0.31-0.92) |
| **KYN** | 1.5 (1-2.3) | 1.6 (1.1-2.7) | 1.5 (1-2.3) | 1.3 (0.9-2.1) | 1.4 (0.91-2.2) | 0.56 (0.29-0.93) | 0.65 (0.35-1) | 0.55 (0.29-0.95) | 0.62 (0.3-1.1) | 0.72 (0.37-1.2) | 1.2 (0.87-1.8) | 1.4 (0.94-2.2) | 1.2 (0.86-1.9) | 1.1 (0.79-1.8) | 1.2 (0.82-1.9) |
| **TRP** | 0.71 (0.52-0.97) | 0.71 (0.52-0.96) | 0.69 (0.5-0.94) | 0.75 (0.54-1) | 0.73 (0.52-0.99) | 0.83 (0.55-1.2) | 0.85 (0.56-1.3) | 0.8 (0.52-1.2) | 0.99 (0.64-1.6) | 1 (0.62-1.6) | 0.76 (0.55-1.1) | 0.76 (0.55-1.1) | 0.76 (0.54-1) | 0.78 (0.55-1.1) | 0.77 (0.54-1.1) |
| **C4-carnitine** | 1.1 (0.81-1.6) | 1.1 (0.84-1.6) | 1.1 (0.81-1.6) | 1 (0.74-1.5) | 1 (0.72-1.5) | 0.54 (0.21-1) | 0.57 (0.22-1.1) | 0.53 (0.2-1) | 0.62 (0.24-1.2) | 0.66 (0.25-1.3) | 0.97 (0.69-1.4) | 0.99 (0.71-1.4) | 0.95 (0.68-1.3) | 0.89 (0.61-1.3) | 0.9 (0.61-1.3) |
| **iso-C4-carnitine** | 1.3 (0.91-1.9) | 1.3 (0.93-2) | 1.2 (0.89-1.9) | 1.2 (0.83-1.8) | 1.2 (0.79-1.8) | 0.55 (0.2-1.1) | 0.63 (0.25-1.1) | 0.56 (0.2-1) | 0.62 (0.21-1.3) | 0.72 (0.28-1.4) | 1 (0.75-1.5) | 1.1 (0.77-1.6) | 1 (0.74-1.5) | 0.97 (0.7-1.5) | 0.98 (0.69-1.5) |
| **ddhC** | 65 (18-348) | 70 (18-369) | 70 (18-350) | 52 (13-289) | 53 (13-273) | 22 (6.4-102) | 21 (6.4-106) | 23 (6.8-107) | 17 (5.4-85) | 17 (5.2-79) | 37 (11-192) | 34 (9.9-157) | 36 (11-180) | 28 (8.6-134) | 30 (8.9-154) |
| **KYN/TRP** | 1.1 (0.81-1.6) | 1.1 (0.81-1.7) | 1.1 (0.81-1.6) | 1 (0.73-1.4) | 1 (0.71-1.4) | 0.35 (0.082-0.85) | 0.43 (0.11-0.9) | 0.34 (0.083- | 0.3 (0.067- | 0.39 (0.091-0.93) | 0.94 (0.68-1.3) | 0.97 (0.69-1.4) | 0.94 (0.68-1.3) | 0.9 (0.61-1.3) | 0.9 (0.63-1.3) |
| **KA/KYN** | 0.67 (0.42-0.96) | 0.68 (0.41-0.96) | 0.63 (0.38-0.92) | 0.64 (0.37-0.95) | 0.6 (0.34-0.93) | 0.37 (0.095-0.89) | 0.33 (0.068- | 0.34 (0.074- | 0.39 (0.097- | 0.35 (0.075-0.9) | 0.38 (0.14-0.76) | 0.38 (0.15-0.76) | 0.34 (0.13-0.71) | 0.38 (0.14-0.77) | 0.33 (0.11-0.72) |
| **KA/TRP** | 0.79 (0.55-1.1) | 0.8 (0.56-1.1) | 0.78 (0.55-1.1) | 0.75 (0.51-1.1) | 0.73 (0.49-1) | 0.12 (0.0094-0.61) | 0.14 (0.011- | 0.12 (0.01-0.61) | 0.13 (0.0096- | 0.16 (0.011-0.7) | 0.58 (0.29-0.91) | 0.58 (0.31-0.9) | 0.55 (0.27-0.88) | 0.55 (0.26-0.89) | 0.53 (0.25-0.89) |
| **C4/Iso-C4 carnitine** | 1.1 (0.82-1.8) | 1.1 (0.83-1.7) | 1.1 (0.83-1.8) | 1.1 (0.83-1.8) | 1.2 (0.81-1.8) | 1.4 (0.85-2.8) | 1.4 (0.86-2.8) | 1.4 (0.86-2.8) | 1.4 (0.82-2.8) | 1.3 (0.8-2.8) | 1.1 (0.8-1.7) | 1.1 (0.76-1.7) | 1.1 (0.78-1.8) | 1.1 (0.77-1.7) | 1.1 (0.74-1.7) |

*SI Table 8. Logistic regression results corrected for age, sex and BMI in COVID-19 severe cases compared to non-severe and deceased compared to discharged patients.*

| | Mild & Intermediate COVID vs Severe COVID | | | | | Discharged vs. Deceases | | | | |
|---|---|---|---|---|---|---|---|---|---|---|
| | **No correction** | **Age** | **Sex** | **BMI** | **Age, Sex & BMI** | **No correction** | **Age** | **Sex** | **BMI** | **Age, Sex & BMI** |
| **KYNA** | 3.4 (1.7-9.2) | 2.9 (1.5-7.7) | 3.4 (1.7-8.8) | 2.7 (1.4-6.3) | 1.9 (1.1-4.6) | 2.3 (1.5-4) | 1.8 (1.2-3.1) | 2.3 (1.5-4.1) | 2.3 (1.5-4) | 1.8 (1.1-3.1) |
| **KYN** | 3.1 (1.8-5.9) | 2.9 (1.6-5.8) | 3.2 (1.9-6.1) | 2.2 (1.3-4.1) | 1.5 (0.84-3) | 1.6 (1.1-2.7) | 1.2 (0.82-2) | 1.6 (1.1-2.7) | 1.5 (1-2.5) | 1.1 (0.68-1.7) |
| **TRP** | 0.78 (0.55-1.1) | 0.81 (0.56-1.2) | 0.78 (0.54-1.1) | 0.78 (0.51-1.1) | 0.78 (0.5-1.2) | 0.68 (0.43-1) | 0.7 (0.44-1.1) | 0.67 (0.44-1) | 0.68 (0.43-1) | 0.68 (0.43-1.1) |
| **C4-Carnitine** | 1.9 (1.3-3.1) | 1.7 (1.1-2.8) | 1.9 (1.3-3.1) | 1.6 (1.1-2.6) | 1.4 (0.89-2.3) | 1.7 (1.2-2.6) | 1.5 (0.99-2.3) | 1.7 (1.1-2.6) | 1.6 (1.1-2.4) | 1.3 (0.84-2) |
| **Iso-C4-carnitine** | 2.6 (1.6-4.4) | 2.3 (1.4-4.2) | 2.6 (1.6-4.5) | 2.8 (1.7-5.3) | 2.3 (1.3-4.5) | 1.9 (1.3-2.9) | 1.6 (1.1-2.5) | 1.9 (1.3-3) | 1.9 (1.3-3) | 1.5 (0.99-2.4) |
| **ddhC** | 1.1 (0.75-1.5) | 0.99 (0.69-1.4) | 1.1 (0.76-1.6) | 0.89 (0.58-1.3) | 0.83 (0.53-1.3) | 1.5 (1-2.2) | 1.4 (0.9-2.1) | 1.6 (1-2.4) | 1.5 (0.97-2.2) | 1.5 (0.94-2.4) |
| **KYN/TRP** | 2.8 (1.7-5.4) | 2.6 (1.5-4.9) | 2.8 (1.8-5.3) | 2 (1.3-3.6) | 1.5 (0.9-2.8) | 2 (1.3-3.1) | 1.6 (1.1-2.5) | 2 (1.3-3.1) | 1.9 (1.3-3) | 1.5 (0.93-2.3) |
| **KYNA/KYN** | 2 (1.3-3.7) | 1.9 (1.1-3.4) | 2 (1.3-3.7) | 2.6 (1.4-5.8) | 2.3 (1.2-4.7) | 2.7 (1.6-5) | 2.4 (1.4-4.5) | 2.8 (1.7-5.3) | 2.9 (1.7-5.6) | 2.6 (1.5-5.2) |
| **KA/TRP** | 3.7 (1.7-12) | 3.1 (1.4-9.9) | 3.9 (1.7-13) | 3 (1.4-8.1) | 2 (1-5.6) | 2.7 (1.6-5.9) | 2.1 (1.2-4.4) | 2.8 (1.5-6.1) | 2.6 (1.5-5.7) | 2 (1.2-4.3) |
| **C4/Iso-C4 carnitine** | 0.94 (0.64-1.3) | 0.98 (0.65-1.5) | 0.95 (0.63-1.4) | 1 (0.65-1.6) | 1.1 (0.7-1.6) | 1 (0.66-1.5) | 1.1 (0.7-1.6) | 1 (0.65-1.5) | 1 (0.64-1.5) | 1.1 (0.69-1.7) |

#### 3.2.2 Outcome in severe COVID-19 cases

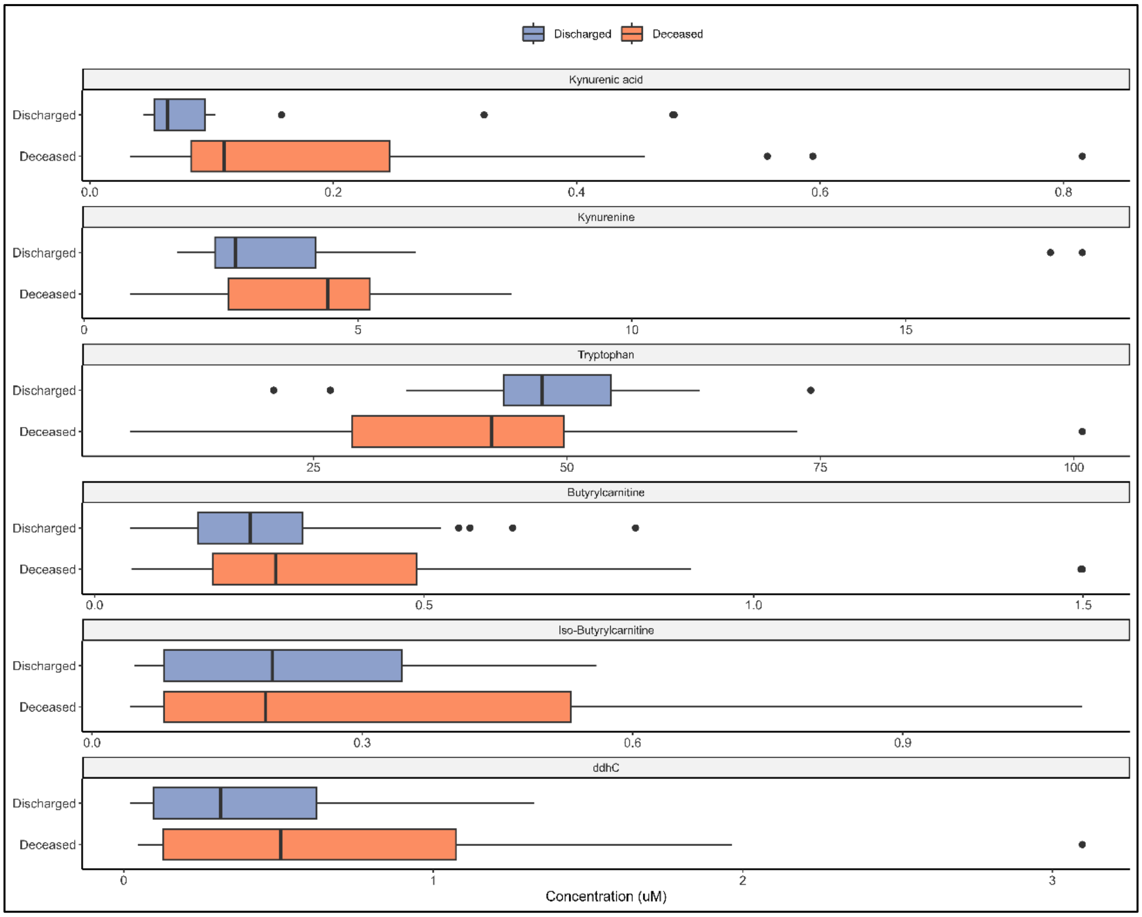


*SI Figure 10. Compound concentrations for outcome in severe COVID patients. Boxes represent the quartiles Q1 to Q3 with Q2 (i.e., median) line in the middle. The 'whiskers' depict the upper and lower limit i.e., Q1 ± (Q3-Q1). Outliers are represented in black circles.*

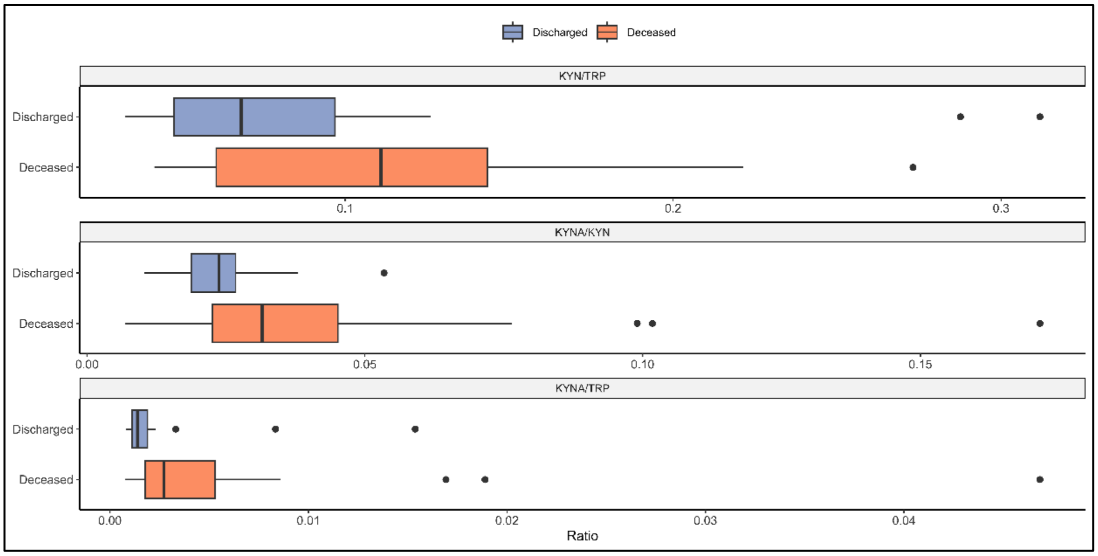


*SI Figure 11. Compound ratios of outcome in severe COVID patients. Boxes represent the quartiles Q1 to Q3 with Q2 (i.e., median) line in the middle. The 'whiskers' depict the upper and lower limit i.e., Q1 ± (Q3-Q1). Outliers are represented in black circles.*

*SI Table 9. Logistic regression results on outcome divergence in severe COVID patients. All results are presented as OR and (90% CI). Significance is established based on CI not crossing 1. Significant relationships with OR > 2 or < 0.5 are coloured in dark blue and significant relationship with smaller effect are coloured in light blue to support readability.*

| Compound | Outcome in severe cases (Deceased vs Discharged) | | | |
|---|---|---|---|---|
| | | Corrected for | | |
| | | Age | Sex | Age & Sex |
| **KYNA** | 2.1 (1.1-5.2) | 1.7 (0.87-4.2) | 2.2 (1.1-5.4) | 1.7 (0.87-4.1) |
| **KYN** | 1 (0.59-1.9) | 0.8 (0.4-1.5) | 1 (0.59-1.8) | 0.79 (0.39-1.5) |
| **TRP** | 0.7 (0.39-1.2) | 0.65 (0.36-1.1) | 0.67 (0.38-1.2) | 0.61 (0.34-1.1) |
| **C4-carnitine** | 1.5 (0.85-2.9) | 1.3 (0.73-2.7) | 1.5 (0.82-2.9) | 1.3 (0.73-2.6) |
| **Iso-C4-carnitine** | 1.4 (0.82-2.6) | 1.2 (0.64-2.2) | 1.4 (0.79-2.5) | 1.1 (0.63-2) |
| **ddhC** | 2 (1.1-4.1) | 2.1 (1.1-4.7) | 2.4 (1.2-5.2) | 2.5 (1.2-5.9) |
| **KYN/TRP** | 1.5 (0.88-2.8) | 1.2 (0.71-2.4) | 1.5 (0.84-2.7) | 1.2 (0.7-2.4) |
| **KYNA/KYN** | 4.6 (1.7-20) | 3.2 (1.3-13) | 4.7 (1.7-20) | 3.6 (1.3-14) |
| **KYNA/TRP** | 3.2 (1.2-14) | 2.4 (1-9.1) | 3.3 (1.3-13) | 2.5 (1-9.9) |
| **C4/Iso-C4-carnitine** | 1.2 (0.72-2.4) | 1.7 (0.85-4.3) | 1.3 (0.69-2.6) | 1.7 (0.83-4.5) |

##### 3.2.3 Compounds and clinical data correlations

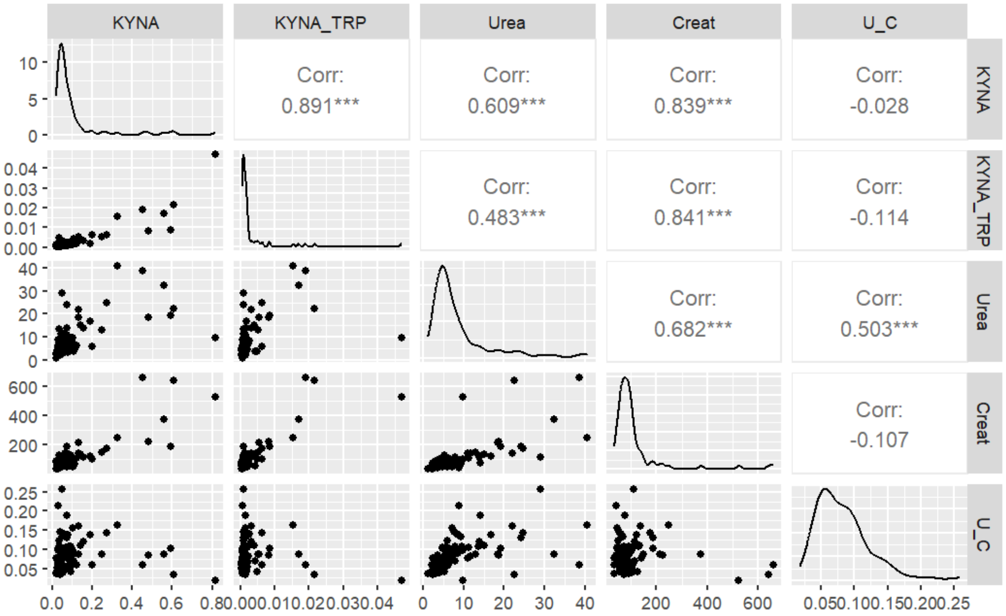


*Figure 12. KYNA and KYNA/TRP correlation to urea, creatinine, and urea/creatinine (U_C) ratio. The strongest correlation is found with creatinine, but no correlation with urea/creatinine ratio.*

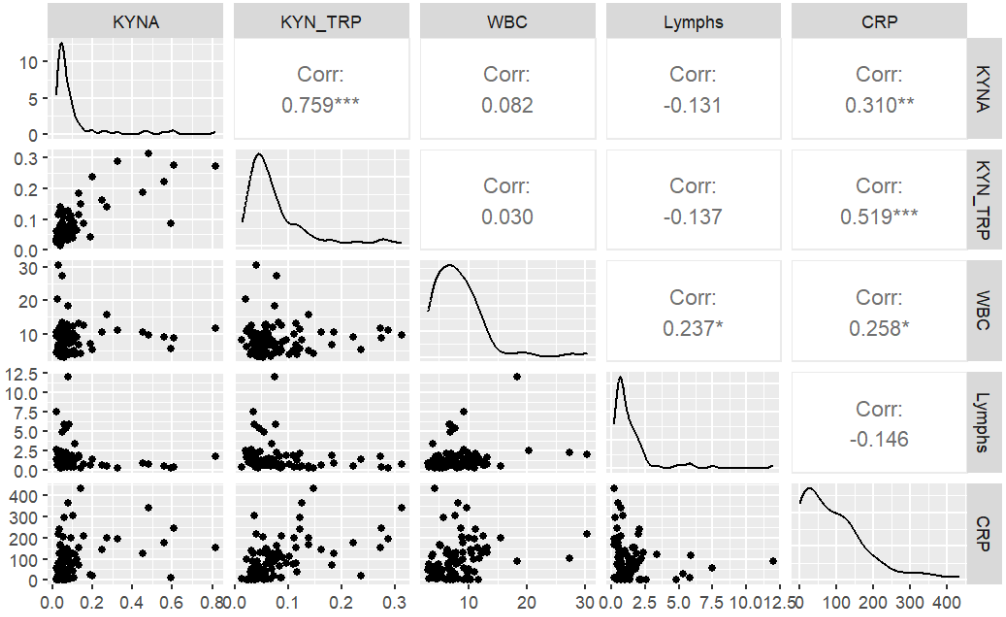


*Figure 13. KYNA and KYNA/TRP correlation to white blood cell count (WBC), lymphocytes count and C-reactive protein (CRP) levels. KYNA/TRP ratio shows stronger correlation to CRP than KYNA alone. No correlation is found to WBC or Lymphs.*

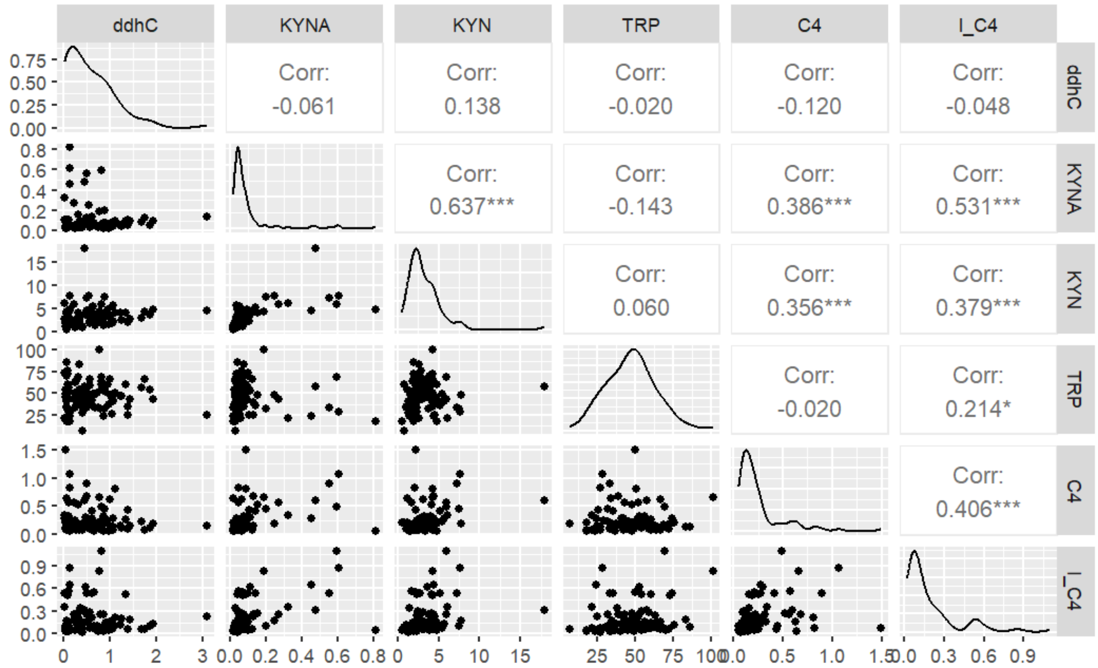


*Figure 14. Correlation analysis between the 6 compounds quantitated in this study. The strongest correlation can be seen between KYNA and KYN, KYNA and iso-C4-carnitine. ddhC is not correlated with the other compounds.*

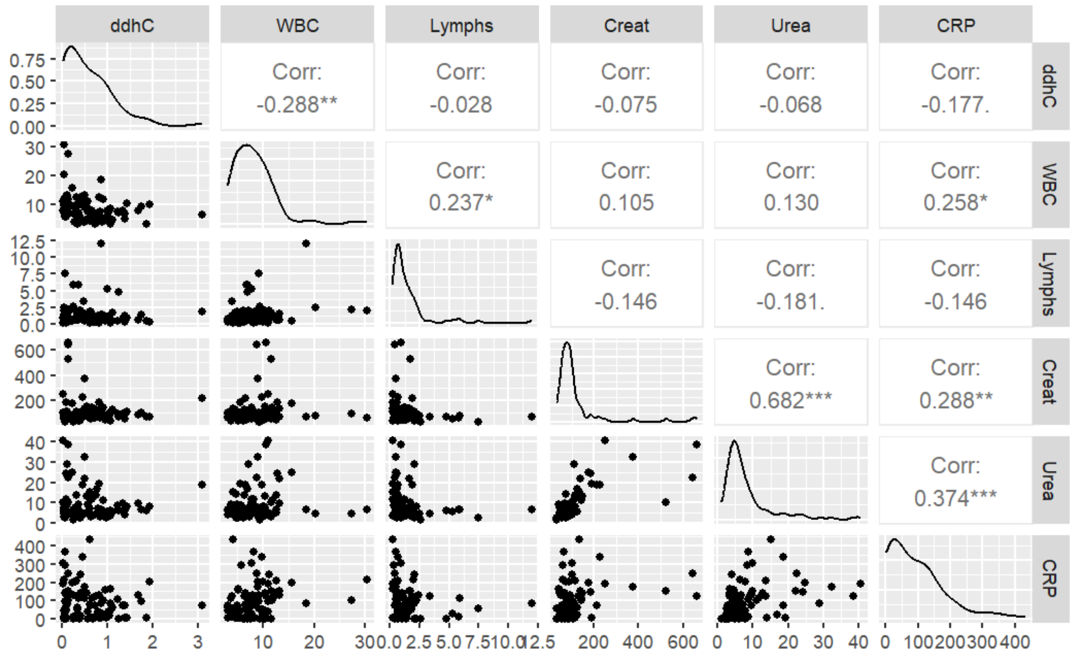


*Figure 20. ddhC correlation exploration to clinical data considered in this study. ddhC is not correlated with the selected clinal markers.*

**Appendix A**

Sample injection order per plate.

| File Name | Position |
|---|---|
| Blank_01 | G:A12 |
| Blank_02 | G:A12 |
| Blank_istd_01 | G:A11 |
| Blank_istd_02 | G:A11 |
| BCQC_01 | B:F8 |
| BCQC_02 | B:F8 |
| BCQC_03 | B:F8 |
| BCQC_04 | B:F8 |
| BCQC_05 | G:B1 |
| BC0_01 | G:A1 |
| BC9_01 | G:A2 |
| BC8_01 | G:A3 |
| BC7_01 | G:A4 |
| BC6_01 | G:A5 |
| BC5_01 | G:A6 |
| BC4_01 | G:A7 |
| BC3_01 | G:A8 |
| BC2_01 | G:A9 |
| BC1_01 | G:A10 |
| BCQC_06 | G:B1 |
| BQC_none_01 | G:B3 |
| BQC_C9_01 | G:B4 |
| BQC_C8_01 | G:B5 |
| BQC_C7_01 | G:B6 |
| BQC_C6_01 | G:B7 |
| BQC_C5_01 | G:B8 |
| BQC_C4_01 | G:B9 |
| BQC_C3_01 | G:B10 |
| BQC_C2_01 | G:B11 |
| BQC_C1_01 | G:B12 |
| BCQC_07 | G:B1 |
| BC0_rep_01 | G:G1 |
| BC9_rep_01 | G:G2 |
| BC8_rep_01 | G:G3 |
| BC7_rep_01 | G:G4 |
| BC6_rep_01 | G:G5 |
| BC5_rep_01 | G:G6 |
| BC4_rep_01 | G:G7 |
| BC3_rep_01 | G:G8 |
| BC2_rep_01 | G:G9 |

| | |
|---|---|
| BC1_rep_01 | G:G10 |
| BCQC_08 | G:B1 |
| BQC_C9_rep_01 | G:H1 |
| BQC_C8_rep_01 | G:H2 |
| BQC_C7_rep_01 | G:H3 |
| BQC_C5_rep_01 | G:H4 |
| BQC_C3_rep_01 | G:H5 |
| BQC_C2_rep_01 | G:H6 |
| BCQC_09 | G:B1 |
| BC0_02 | G:A1 |
| BC9_02 | G:A2 |
| BC8_02 | G:A3 |
| BC7_02 | G:A4 |
| BC6_02 | G:A5 |
| BC5_02 | G:A6 |
| BC4_02 | G:A7 |
| BC3_02 | G:A8 |
| BC2_02 | G:A9 |
| BC1_02 | G:A10 |
| BCQC_06 | G:B1 |
| BQC_none_02 | G:B3 |
| BQC_C9_02 | G:B4 |
| BQC_C8_02 | G:B5 |
| BQC_C7_02 | G:B6 |
| BQC_C6_02 | G:B7 |
| BQC_C5_02 | G:B8 |
| BQC_C4_02 | G:B9 |
| BQC_C3_02 | G:B10 |
| BQC_C2_02 | G:B11 |
| BQC_C1_02 | G:B12 |
| SCQC_01 | G:C1 |
| SCQC_02 | G:C1 |
| SCQC_03 | G:C1 |
| SCQC_04 | G:C2 |
| SQC_none_01 | G:C2 |
| SQC_C9_01 | G:C3 |
| SQC_C8_01 | G:C4 |
| SQC_C7_01 | G:C5 |
| SQC_C6_01 | G:C6 |
| SQC_C5_01 | G:C7 |
| SQC_C4_01 | G:C8 |
| SQC_C3_01 | G:C9 |
| SQC_C2_01 | G:C10 |
| SQC_C1_01 | G:C11 |

| | |
|---|---|
| SQC_C9_rep_01 | G:H7 |
| SQC_C8_rep_01 | G:H8 |
| SQC_C7_rep_01 | G:H9 |
| SQC_C5_rep_01 | G:H10 |
| SQC_C3_rep_01 | G:H11 |
| SQC_C2_rep_01 | G:H12 |
| Plate_QC_01 | G:G11 |
| Plate_QC_02 | G:G11 |
| Plate_QC_03 | G:G11 |
| Plate_QC_04 | G:G11 |
| Study_QC_01 | G:C12 |
| S_03_01 | G:D3 |
| S_28_01 | G:F4 |
| S_35_01 | G:F11 |
| S_13_01 | G:E1 |
| S_07_01 | G:D7 |
| S_31_01 | G:F7 |
| Plate_QC_05 | G:G11 |
| S_09_01 | G:D9 |
| S_14_01 | G:E2 |
| S_15_01 | G:E3 |
| S_10_01 | G:D10 |
| S_36_01 | G:F12 |
| S_20_01 | G:E8 |
| Study_QC_02 | G:C12 |
| Plate_QC_06 | G:G11 |
| S_34_01 | G:F10 |
| S_29_01 | G:F5 |
| S_12_01 | G:D12 |
| S_24_01 | G:E12 |
| S_25_01 | G:F1 |
| S_22_01 | G:E10 |
| Plate_QC_07 | G:G12 |
| S_08_01 | G:D8 |
| S_19_01 | G:E7 |
| S_06_01 | G:D6 |
| S_23_01 | G:E11 |
| S_33_01 | G:F9 |
| S_04_01 | G:D4 |
| Study_QC_03 | G:C12 |
| Plate_QC_08 | G:G12 |
| S_30_01 | G:F6 |
| S_05_01 | G:D5 |
| S_18_01 | G:E6 |

| | |
|---|---|
| S_17_01 | G:E5 |
| S_01_01 | G:D1 |
| S_21_01 | G:E9 |
| Plate_QC_09 | G:G12 |
| S_27_01 | G:F3 |
| S_02_01 | G:D2 |
| S_32_01 | G:F8 |
| S_11_01 | G:D11 |
| S_26_01 | G:F2 |
| S_16_01 | G:E4 |
| Study_QC_04 | G:C12 |
| Plate_QC_10 | G:G12 |
| S_06_02 | G:D6 |
| S_18_02 | G:E6 |
| S_26_02 | G:F2 |
| S_15_02 | G:E3 |
| S_03_02 | G:D3 |
| S_21_02 | G:E9 |
| Plate_QC_11 | G:G12 |
| SCQC_04 | G:C1 |
| SCQC_05 | G:C1 |
| SCQC_06 | G:C1 |
| SQC_none_02 | G:C2 |
| SQC_C9_02 | G:C3 |
| SQC_C8_02 | G:C4 |
| SQC_C7_02 | G:C5 |
| SQC_C6_02 | G:C6 |
| SQC_C5_02 | G:C7 |
| SQC_C4_02 | G:C8 |
| SQC_C3_02 | G:C9 |
| SQC_C2_02 | G:C10 |
| SQC_C1_02 | G:C11 |
| SQC_C9_rep_02 | G:H7 |
| SQC_C8_rep_02 | G:H8 |
| SQC_C7_rep_02 | G:H9 |
| SQC_C5_rep_02 | G:H10 |
| SQC_C3_rep_02 | G:H11 |
| SQC_C2_rep_02 | G:H12 |
| BCQC_07 | B:F8 |
| BCQC_08 | B:F8 |
| BCQC_09 | G:B2 |
| BC0_03 | G:A1 |
| BC9_03 | G:A2 |
| BC8_03 | G:A3 |

| | |
|---|---|
| BC7_03 | G:A4 |
| BC6_03 | G:A5 |
| BC5_03 | G:A6 |
| BC4_03 | G:A7 |
| BC3_03 | G:A8 |
| BC2_03 | G:A9 |
| BC1_03 | G:A10 |
| BCQC_10 | G:B2 |
| BQC_none_03 | G:B3 |
| BQC_C9_03 | G:B4 |
| BQC_C8_03 | G:B5 |
| BQC_C7_03 | G:B6 |
| BQC_C6_03 | G:B7 |
| BQC_C5_03 | G:B8 |
| BQC_C4_03 | G:B9 |
| BQC_C3_03 | G:B10 |
| BQC_C2_03 | G:B11 |
| BQC_C1_03 | G:B12 |
| BCQC_11 | G:B2 |
| BC0_rep_02 | G:G1 |
| BC9_rep_02 | G:G2 |
| BC8_rep_02 | G:G3 |
| BC7_rep_02 | G:G4 |
| BC6_rep_02 | G:G5 |
| BC5_rep_02 | G:G6 |
| BC4_rep_02 | G:G7 |
| BC3_rep_02 | G:G8 |
| BC2_rep_02 | G:G9 |
| BC1_rep_02 | G:G10 |
| BCQC_12 | G:B2 |
| BQC_C9_rep_02 | G:H1 |
| BQC_C8_rep_02 | G:H2 |
| BQC_C7_rep_02 | G:H3 |
| BQC_C5_rep_02 | G:H4 |
| BQC_C3_rep_02 | G:H5 |
| BQC_C2_rep_02 | G:H6 |
| Blank_istd_03 | G:A11 |
| Blank_istd_04 | G:A11 |
| Blank_03 | G:A12 |
| Blank_04 | G:A12 |
